# Unraveling the relationship of loneliness and isolation in schizophrenia: Polygenic dissection and causal inference

**DOI:** 10.1101/2020.11.06.20226910

**Authors:** A. Andreu-Bernabeu, C.M. Díaz-Caneja, J. Costas, L. de Hoyos, C. Stella, X. Gurriarán, C. Alloza, L. Fañanás, J. Bobes, A. González Pinto, B. Crespo-Facorro, L. Martorell, E. Vilella, G. Muntane, J. Nacher, M.D. Molto, E.J. Aguilar, M. Parellada, C. Arango, J. González-Peñas

## Abstract

There is increasing recognition of the association between loneliness and social isolation (LNL-ISO) with schizophrenia. Here, we demonstrate significant LNL-ISO polygenic score prediction on schizophrenia in an independent case-control sample (N=3,488). We then dissect schizophrenia predisposing variation into subsets of variants based on their effect on LNL-ISO. Genetic variation with concordant effects in both phenotypes show significant SNP-based heritability enrichment, higher polygenic predictive ability in females and positive covariance with other mental disorders such as depression, anxiety, attention-deficit hyperactivity, alcohol use disorder, and autism. Conversely, genetic variation with discordant effects is only predictive in males and negatively correlated with those disorders. This correlation pattern is not observed for bipolar and obsessive-compulsive disorders. Mendelian randomization analyses demonstrate a plausible bi-directional causal relationship between LNL-ISO and schizophrenia, with a greater effect of LNL-ISO liability on schizophrenia. These results illustrate the genetic footprint of LNL-ISO on schizophrenia and suggest its role as a potential target for early intervention.

## BACKGROUND

Social relationships are critical for emotional and cognitive development in social species such as humans ^1,2^. In fact, the need to belong to a social group has been classically posed as a fundamental behavior in our species ^3^. Both objective and perceived (i.e., loneliness) social isolation have been characterized ^4,5^. While the former may be defined as an objective lack of social connections (interactions, contacts or relationships), the latter refers to the subjective feeling of distress associated with a lack of meaningful relationships, regardless of the amount of social contacts ^6^. Although isolated people often feel lonely, both entities are not always correlated ^4–6^. However, regardless of type, both objective social isolation and loneliness have been recognized as major risk factors for morbidity and mortality ^6–8^, as well as for the onset of mental disorders ^9–14^.

Although loneliness and objective social isolation have been typically associated with depressive symptoms and major depression ^14–16^, a renewed interest has grown in the study of their association with psychosis ^17–20^. Social withdrawal and isolation are described in early stages of schizophrenia ^17,21,22^, recalling the classical descriptions of pre-schizophrenia related traits from psychiatrists such as Kraepelin, Bleuler or Conrad ^23–25^. Indeed, recent meta-analyses indicate that loneliness plays an important role in the onset and maintenance of psychotic symptoms ^17,22,26^. Another meta-analysis also showed a consistent association of loneliness with both positive and negative psychotic-like experiences ^27^. Moreover, there are studies suggesting that loneliness may increase subclinical paranoia in non-clinical populations ^28^. However, the causal relationships between social isolation and schizophrenia are still unclear ^17,29^.

Inherited biological factors could explain, at least partially, the relationship between social isolation and schizophrenia. Available evidence supports the genetic basis of loneliness and objective social isolation ^30,5,31^. A recent study used multi-trait GWAS (MTAG) ^32^, a software developed to jointly analyze different summary statistics from related traits, to assess the genetic architecture of loneliness and objective social isolation (LNL-ISO) by combining the three UK Biobank GWAS datasets of i) perceived loneliness, ii) a proxy of social support (combined frequency of family/friends visits and living alone), and iii) ability to confide in someone close ^33^. Up to 15 genome-wide significant *loci* and SNP-based heritability estimates (*h*^*2*^_*SNP*_ = 4.2%) support the contribution of common genetic variation to this social construct. This study also reported a significant genetic correlation of this combined phenotype (LNL-ISO) with schizophrenia, along the lines of a previous study reporting a significant association of perceived loneliness with schizophrenia, but not with bipolar disorder^30^. Schizophrenia polygenic scores have also been found to significantly predict loneliness in an independent population, thus lending further support to a shared genetic etiology between both phenotypes ^34^.

Previous studies exploring the genetic relationship between perceived and objective social isolation and schizophrenia leave several questions unanswered, including the direction of their association, the specific biological effects of shared and non-shared predisposing variants, and the effect of additional factors on this relationship, including sex. The epidemiological and clinical presentation of psychotic disorders differs between sexes ^35–37^ and sex also seems to affect the perception of loneliness and the psychological impact of isolation, although results have been contradictory so far ^37–40^.

We aimed to test the hypothesis that there is a bidirectional genetic relationship between perceived and objective social isolation and schizophrenia within a systematic and comprehensive framework. First, we analyzed loneliness and social isolation (LNL-ISO) polygenic score prediction on schizophrenia risk in an independent Spanish case-control sample. Second, we dissected the predisposing variation to schizophrenia according to its role in LNL-ISO, and analyzed the polygenic risk predictions, biological profiles (using brain specific functional annotations), and sex effects across each genomic partition using a novel approach. Third, to evaluate the role of LNL-ISO in the genetic overlap between psychiatric disorders and other related traits, we studied the partial correlations between schizophrenia and related phenotypes across the aforedescribed LNL-ISO partitions. Finally, we performed a causality analysis between LNL-ISO and schizophrenia using a two-sample Mendelian randomization approach.

## METHODS

### Samples and GWAS summary data

A case-control sample including 1927 schizophrenia cases (65% males) and 1561 healthy controls (HC) (55% males) from CIBERSAM (Centro de Investigación Biomédica en Red en Salud Mental, Spain) was used as an independent target sample for polygenic scores predictions (SCZ_CIBERSAM). All participants were genotyped as part of the Psychiatric Genomics Consortium (PGC), and passed quality control (QC filters) per PGC-SZ2 criteria ^41^. See **Supplementary Methods** for a detailed description.

We used the following genetic summary statistics from previous GWAS: i) schizophrenia GWAS from the Psychiatric Genetic Consortium (PGC-SCZ2) comprising 35,476 cases and 46,839 controls ^41^, ii) loneliness and social isolation combined phenotype (LNL-ISO)^33^ GWAS based on the combined multi-trait GWAS (MTAG) in the UK Biobank (UKBB) study, yielding an effective maximum sample size of 487,647 individuals, and iii) the latest UKBB GWAS results for the independent loneliness and isolation traits that were included in the original LNL-ISO MTAG: a) *Loneliness UKBB* (https://nealelab.github.io/UKBB_ldsc/h2_summary_2020.html), b) a proxy of social support, as measured by the frequency *of family and friend visits* (https://nealelab.github.io/UKBB_ldsc/h2_summary_1031.html) and the *number of people living in household* (https://nealelab.github.io/UKBB_ldsc/h2_summary_709.html) and c) *ability to confide* (https://nealelab.github.io/UKBB_ldsc/h2_summary_2110.html).

### Dissection of schizophrenia summary genetic data based on LNL-ISO

First, we selected variants that were included in both schizophrenia ^41^ and LNL-ISO summary data. Second, we divided schizophrenia summary statistics from the set of overlapping variants (N_SNPs = 5,658,282) into two different subsets of variants, according to their effects in the LNL-ISO: those variants not associated with LNL-ISO (SCZ_noLNL; P_LNL-ISO_ > 0.05; N_SNPs = 5,172,017) and those variants associated with LNL-ISO (**SCZ_LNL**; P_LNL-ISO_ < 0.05; N_SNPs = 486,265). This cutoff was selected based on the fact that LNL-ISO-based PGS (PGS_LNL-ISO_) prediction on schizophrenia in the case-control target sample from CIBERSAM at P > 0.05 was not significant (R^2^ =0.0031% at *P*_*threshold*_ = 0.05 - 1, *P* = 0.768; **Supplementary Table 1 C)**. Third, based on the concordance or discordance of the effects of the same effect allele, SCZ_LNL was again divided into those variants with concordant (**SCZ_LNL_CONC**; N_SNPs = 269,361) or discordant (**SCZ_LNL_DISC**; N_SNPs = 216,904) effects in schizophrenia and LNL-ISO (SCZ_LNL_CONC: Beta_SCZ_ > 0 & Beta_LNL-ISO_ > 0 OR Beta_SCZ_ < 0 & Beta_LNL-ISO_ < 0; SCZ_LNL_DISC: Beta_SCZ_ > 0 & Beta_LNL-ISO_ < 0 OR Beta_SCZ_ < 0 & Beta_LNL-ISO_ > 0). In each of the final datasets, correlated SNPs due to linkage disequilibrium (LD) were removed using PLINK 1.9 clumping algorithm (r^2^ > 0.1; window size = 500 kb). See **Supplementary Methods 3** for further details.

**Table 1.**
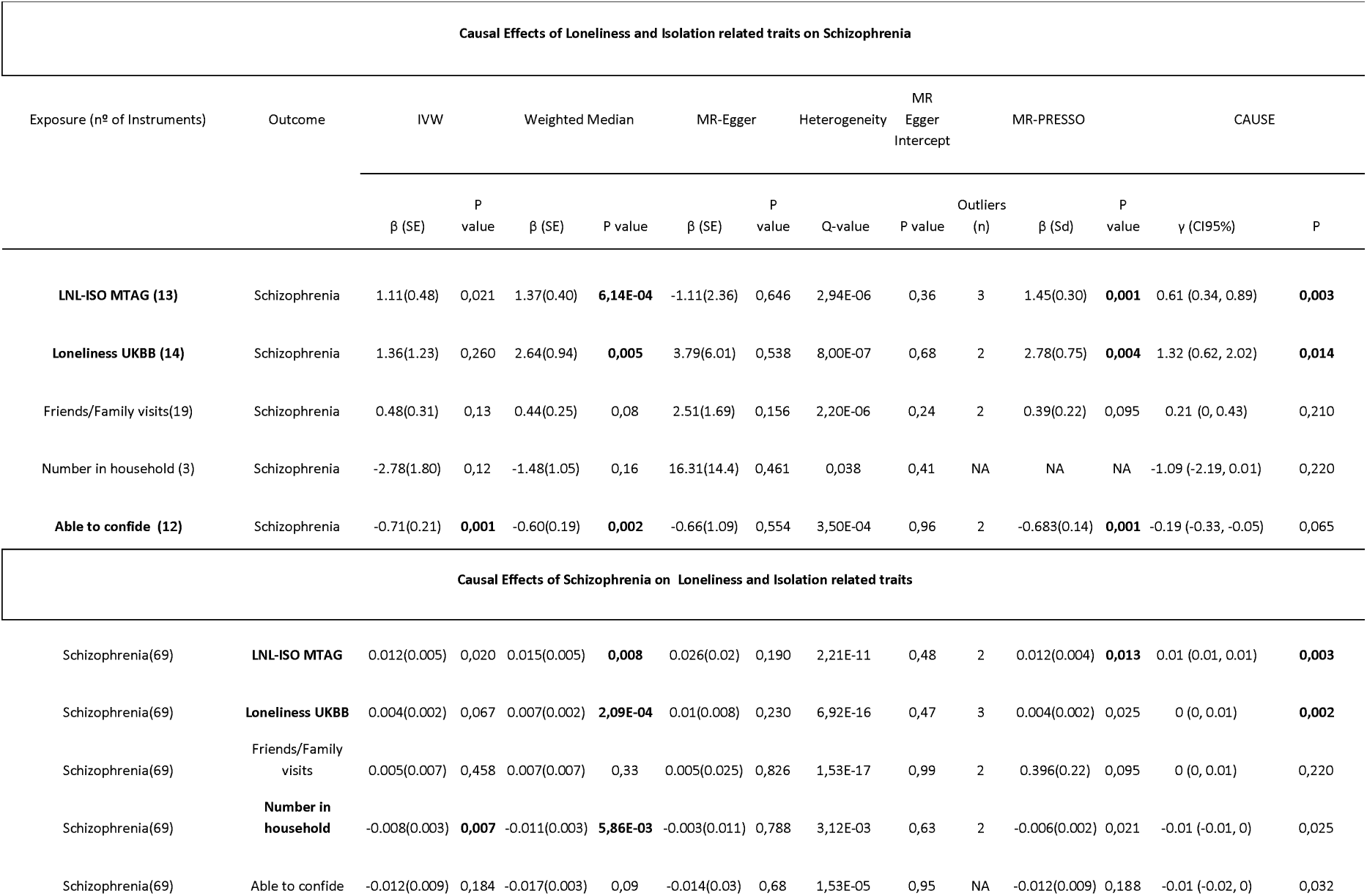
Bidirectional causal inference analyses between Loneliness and Isolation phenotypes and Schizophrenia. Traits in bold have significant results in at least one method after Benjamini-Hochberg FDR correction. P-values labeled in bold are significant after Benjamini-Hochberg FDR correction (P_fdr < 0.05). The column “Outliers” reports the number of pleiotropic variants removed from the MR estimate. Outliers are variants showing possible evidence of horizontal pleiotropy and that were removed in MR-PRESSO before the causal estimate was made. IVW, inverse variance-weighted linear regression. SE, Standard Error measure of effect size. LNL-ISO MTAG, Multitrait GWAS of Loneliness and social isolation. Loneliness UKBB, Loneliness trait from the UK Biobank. Friends/Family visit, UK Biobank trait of Frequency of Friends/Family visits. Able to confide, UK biobank trait of Frequency of confide in someone close to you. γ (CI95%) Posterior median and 95% credible intervals of true value of causal effect with CAUSE. P: p-value testing that causal model is a better fit with CAUSE using ELPD test (significance level p < 0.05)

### Polygenic score (PGS) predictions

Polygenic models were performed based on PGC-SZ2^41^ (PGS) and LNL-ISO ^33^ (PGSLNL_ISO_) GWAS summary statistics as the discovery samples, and SCZ_CIBERSAM case-control sample as the target sample (N_SCZ = 1927; N_HC = 1561). Several P thresholds were used (P<5 × 10^−8^, 5 × 10^−5^, 1 × 10^−3^, 0.01, 0.05, 0.1, 0.2, 0.5 and 1). Standardized PGS were calculated and significance was evaluated by logistic regression, using case-control status as dependent variable and sex, age, and 10 first multidimensional scaling (MDS) ancestry components as covariates. Explained variance attributable to PGS was calculated as the increase in Nagelkerke’s pseudo-R^2^ between a model with and without the PGS variable. All p-values were FDR corrected and predictions at P-threshold that lead to the most significant results were confirmed after 10K permutations of the case-control status (see **Supplementary Figures 1-2**). In order to compare PGS_LNL-ISO_ predictions with LNL-ISO’s constituent phenotypes (*Loneliness UKBB, frequency of family and friend visits, number of people living in household and ability to confide*), their PGS predictions were also evaluated on the same target sample.

**Figure 1.**
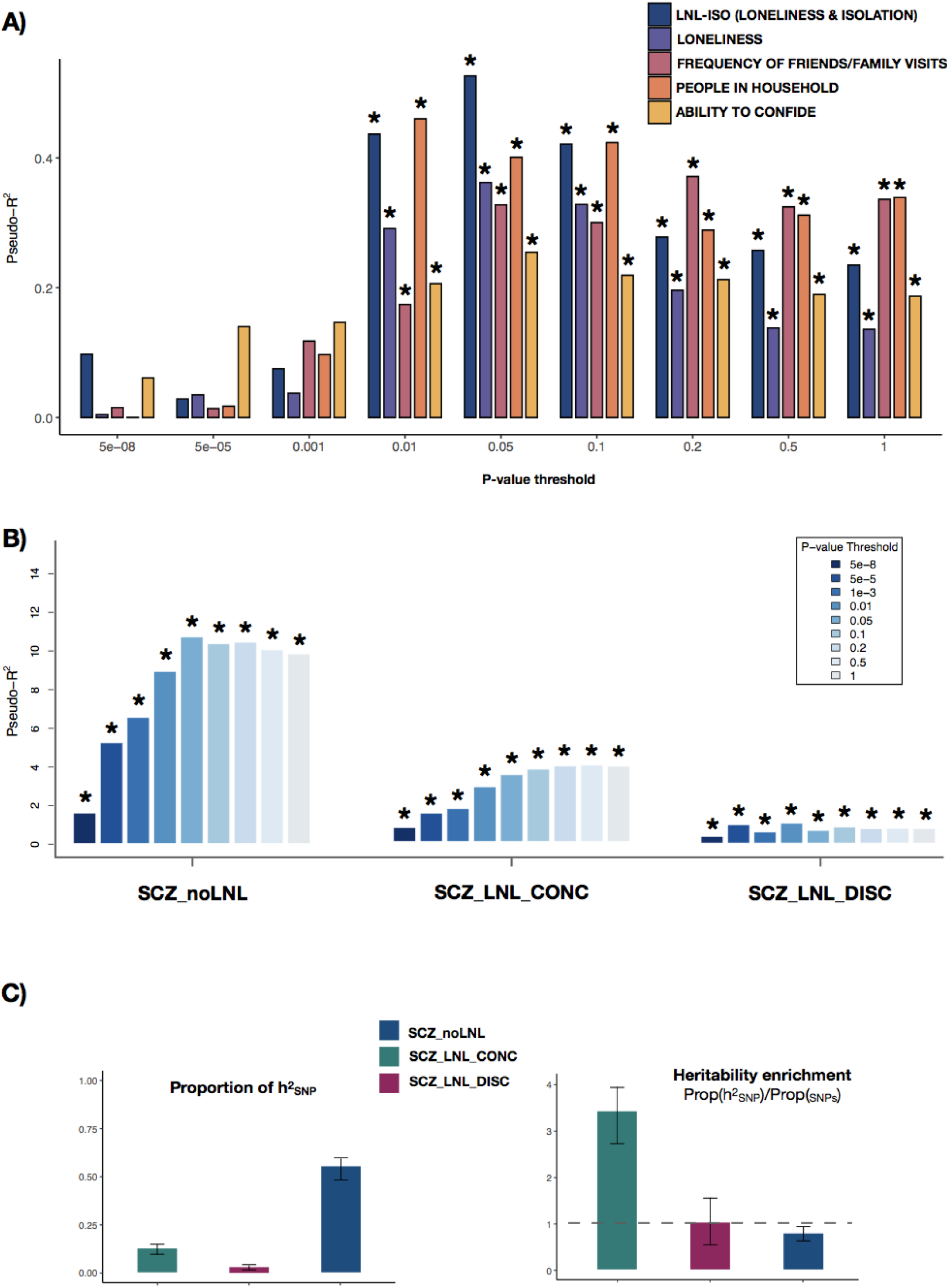
Polygenic score predictions of schizophrenia (PGS_SCZ_) and LNL-ISO (PGS_LNL-ISO_) on schizophrenia and heritability estimates. **A)** PGS predictions of LNL-ISO and its constituent phenotypes (see legend) on an independent schizophrenia case-control sample (N_SCZ_ = 1927; N_HC_ = 1561). Significant tests after FDR correction (*p*_*FDR*_ < 0.05) are marked with an asterisk. **B)** PGS_SCZ_ predictions on an independent schizophrenia case-control sample (N_SCZ_ = 1927; N_HC_ = 1561). SCZ GWAS summary statistics were divided into three groups: those variants not associated with LNL-ISO (**SCZ_noLNL**; *P*_LNL-ISO_ > 0.05), those associated with LNL-ISO, with a concordant sign of the allele effect in both schizophrenia and LNL-ISO (**SCZ_LNL_CONC**; *P*_LNL-ISO_ < 0.05; Beta_SCZ_ > 0 & Beta_LNL-ISO_ > 0 / Beta_SCZ_ < 0 & Beta_LNL-ISO_ < 0) and those associated with LNL-ISO but with a discordant direction of the effect relative to schizophrenia (**SCZ_LNL_DISC**; *P*_LNL-ISO_ < 0.05; Beta_SCZ_ > 0 & Beta_LNL-ISO_ < 0 / Beta_SCZ_ < 0 & Beta_LNL-ISO_ > 0). For full detailed results see **Supplementary Table 2**. C) Proportion of SNP-based heritability (*h*^*2*^_*SNP*_) and heritability enrichment (*h*^*2*^ _*SNP*_/*N*_*SNP*_) of the three LNL-ISO based annotations **(SCZ_noLNL, SCZ_LNL_CONC** and **SCZ_LNL_DISC**) in schizophrenia. 95% confidence intervals based on standard errors are shown for each estimate. See **Supplementary Table 4** for significance for each enrichment estimate.

PGS_SCZ_ were also calculated using the described separated subsets of variants based on their effect in LNL-ISO. LD-independent variants within **SCZ_noLNL** (N_clumped SNPs = 169,574), **SCZ_LNL** (N_clumped SNPs = 11,804), **SCZ_LNL_CONC** (N_clumped SNPs = 6,468) and **SCZ_LNL_DISC** (N_clumped SNPs = 5,336) were used to calculate PGS on the SCZ_CIBERSAM case-control cohort. Standardized PGS were calculated and significance was evaluated by logistic regression models as described above.

In order to assess the effect of sex on these models, we compared the explained variance in the case-control status for predisposing variation to schizophrenia within SCZ_noLNL, SCZ_LNL, SCZ_LNL_CONC and SCZ_LNL_DISC in females and males, by using a permutation approach (5,000 permutations) based on a random selection of 500 schizophrenia and 500 HC subjects in each sex separately (see **Supplementary Methods 3**).

### LD-score regression (LDSR) and heritability estimates

SNP-based heritability (*h*^*2*^_*SNP*_) estimates were calculated for resulting genome partitions from dissections of schizophrenia summary genetic data based on LNL-ISO as described before : i) SCZ_noLNL, SCZ_LNL_CONC, and SCZ_LNL_DISC annotations; and ii) sub-annotations from the intersection between those annotations (SCZ_noLNL and SCZ_LNL_CONC) and gene expression data from 10 whole tissues ^42^, 13 brain-related tissues (Brain GTEx^43^) and 3 brain cell-type annotation files (neurons, astrocytes, and oligodendrocytes ^43,44^). The intersection with SCZ_LNL_DISC was not included due to the low *h*^*2*^_*SNP*_ for this annotation. ldsc v1.0.1 ^43^, a command line tool for estimating heritability, was used. We performed both heritability enrichment analyses across the described annotations (--*h2*) and one-sided t-tests to evaluate whether the cell-type enrichment in schizophrenia within a particular LNL-ISO annotation was higher than the same cell-type enrichment in schizophrenia outside the LNL-ISO annotation (--*h2*-cts) (see **Supplementary Methods 4**).

### Partial genetic correlations between schizophrenia and related phenotypes based on LNL-ISO annotations

To examine the influence of LNL-ISO based annotations (SCZ_noLNL, SCZ_LNL_CONC, and SCZ_LNL_DISC) on the correlation between schizophrenia and other related disorders or traits (**Supplementary Methods 5**), we calculated partial correlations. First, we selected neuropsychiatric and related phenotypes reportedly showing significant correlations with schizophrenia and/or loneliness/social isolation phenotypes in previous studies (i.e., major depression (MDD), attention and deficit hyperactivity disorder (ADHD), autism spectrum disorder (ASD), anxiety disorder (ANX), bipolar disorder (BIP), obsessive compulsive disorder (OCD), alcohol dependence disorder (ALC-DEP), cross-disorder phenotype (CROSS-DIS), neuroticism (NEUR), depressive symptoms (DS), subjective well-being (SWB), psychotic experiences in the general population (PSY-EXP) and educational attainment (EA). We also included body-mass index (BMI) summary data since it has been reported to be strongly influenced by LNL-ISO^33^ (see **Supplementary Methods 5** for references). We used covariance estimates based on partial correlations restricted to SNP subsets within each annotation conducted with GNOVA ^45^. Derived p-values were statistically corrected using a Benjamini-Hochberg False Discovery Rate (FDR) procedure (*p*_*FDR*_ < 0.05).

### Two -sample Mendelian randomization

We used Mendelian Randomization (MR) to investigate the direction of the causal relationships between social isolation (LNL-ISO) and its constituents (i.e. loneliness UKBB, frequency family visits, number of people in household, and ability to confide) with schizophrenia liability using the latest GWAS data available in MRC-IEU API resource^46^.

We used five MR methods - i) Inverse variance weighted method (IVW) ^47,48^, ii) Weighted median (WM)^49^, iii) MR-Eggers^50^, iv) Simple mode (SM), and v) Weighted mode (WMo)^51^- in the R package TwoSampleMR v.0.5.3^52^ with the default settings.

Additionally, we conducted Mendelian Randomization Pleiotropy RESidual Sum and Outlier (MR-PRESSO) ^53^ [https://github.com/rondolab/MR-PRESSO] analyses and a novel MR method (CAUSE) ^54^ to further account for pleiotropy (see **Supplementary Methods 6** for further details).. We applied multiple testing corrections using Benjamini-Hochberg FDR (*p*_*FDR*_ < 0.05)

## RESULTS

### LNL-ISO polygenic score (PGS_LNL-ISO_) prediction on schizophrenia

**Figure 1A** shows the percentage of variance in schizophrenia risk explained by LNL-ISO (PGS_LNL-ISO_) in the CIBERSAM case-control sample. We found that common genetic variation predisposing to LNL-ISO significantly contributed to schizophrenia risk (R^2^ =0.53% at *P*_*threshold*_ = 0.05, *p* = 1.2 × 10^−4^). One standard deviation (s.d.) increase in PGS_LNL-ISO_ was associated with a 15% increase in the likelihood of belonging to the schizophrenia group (OR (95%CI) = 1.15 (1.07 – 1.24)). This contribution remained significant after performing 10,000 case-control status permutations (*p* = 9.95 × 10^−5^; **Supplementary Table 1A, Supplementary Figure 1**). In the same target sample, LNL-ISO explained 46% more variance in schizophrenia risk than loneliness UKBB (R^2^ = 0.36% at *P*_*threshold*_ = 0.05, *p* = 1.42 ×10^−3^; **Figure 1A**). The contribution of PGS_LNL-ISO_ to schizophrenia risk was also higher than that of *ability to confide* (R^2^ =0.25% at *P*_*threshold*_ = 0.05, *p* = 7.4 × 10^−3^) and the two measures of social support included in LNL-ISO: *number of people living in household* (R^2^ =0.46% at *P*_*threshold*_ = 0.01, *p* = 3.14 ×10^−4^) and *frequency of family/friends visits* (R^2^ =0.37% at *P*_*threshold*_ = 1, p = 1.2 ×10^−3^; see **Supplementary table 1B**).

### Polygenic dissection of schizophrenia by its relationship with LNL-ISO

PGS_scz_ predictions on the same schizophrenia case-control sample were performed for the three subsets of SNPs based on the dissection of SCZ summary data according to the role in LNL-ISO: PGS_scz_ predictions from variants only contributing to SCZ (PGS_SCZ_noLNL_) and those contributing to both phenotypes with concordant (PGS_SCZ_LNL_CONC_) and discordant (PGS_SCZ_LNL_DISC_) effects (see **Methods**).

**Figure 1B** shows the percentage of variance in schizophrenia risk explained by PGS_scz_ within each subset of SNPs. PGS_SCZ_LNL_CONC_ explained almost four times more variance (R^2^ = 3.94% at *P*_*threshold*_ = 0.5, *p* = 8.36 × 10^−25^) than PGS SCZ_LNL_DISC (R^2^ = 1.02% at *P*_*threshold*_ = 0.01, p = 8.43 × 10^−8^).

Heritability estimates by LDSR found that variation within **SCZ_LNL_CONC** showed a significant SNP-based heritability (*h*^*2*^_*SNP*_) enrichment, with 3.8% of the SNPs explaining an estimated 13.1% of the SNP-based heritability (Enrichment(CI_95%_) = 3.43 (2.86 - 4.01); *p* = 1.83 × 10^−15^; **Figure 1C; Supplementary Table 4**). By contrast, variants within **SCZ_noLNL** harbored 65% of the SNPs and accounted for around 53.9% of the heritability, with a relative *h*^*2*^_*SNP*_ decrease for this annotation (Enrichment(CI_95%_) = 0.81 (0.72 - 0.90); *p* = 8.12 × 10^−5^; **Figure 1C**). We found no significant heritability enrichment for **SCZ_LNL_DISC** (Enrichment(CI_95%_) = 1.08 (0.58 - 1.58); *p* = 0.748).

Partitioned heritability and LD score regression analyses were also applied to specifically expressed genes (LDSC-SEG) within the described annotations. We observed comparable heritability enrichment profiles were observed for SCZ_noLNL and SCZ_LNL_CONC across the central nervous system (CNS) and the neuronal cell type (**Supplementary Table 4**). Across the 13 brain tissues analyzed, distinct enrichment patterns were observed. Schizophrenia predisposing variation within **SCZ_noLNL** was specifically enriched in GTEx brain cortex (*p* = 8.5 × 10^−4^) and anterior cingulate cortex (*p* = 5.16 × 10^−3^ ; **Supplementary Figure 3**), while predisposing variation within **SCZ_LNL_CONC** was enriched in GTEx hippocampus (*p* = 0.041), although the latter did not passed significance after FDR correction.

We finally assessed PGS_SCZ_ prediction stratified by sex in the CIBERSAM case-control sample based on variation within SCZ_noLNL, SCZ_LNL, SCZ_LNL_CON and SCZ_LNL_DISC (**Figure 2**). PGS_SCZ_LNL_CONC_ explained significantly more variance in schizophrenia risk in females than in males (R^2^ =5.16% at *P*_*threshold*_ = 0.1, *p* = 1.88 × 10^−13^ and R^2^ =3.31% at *P*_*threshold*_ = 0.5, *p* = 2.00 × 10^−13^, respectively), while the opposite pattern was observed for PGS_SCZ_LNL_DISC_ (R^2^ = 0.51% at *P*_*threshold*_ = 0.01, *p* = 0.02 in females and R^2^ =1.68% at *P*_*threshold*_ = 0.1, *p* = 1.55 × 10^−7^ in males).

**Figure 2.**
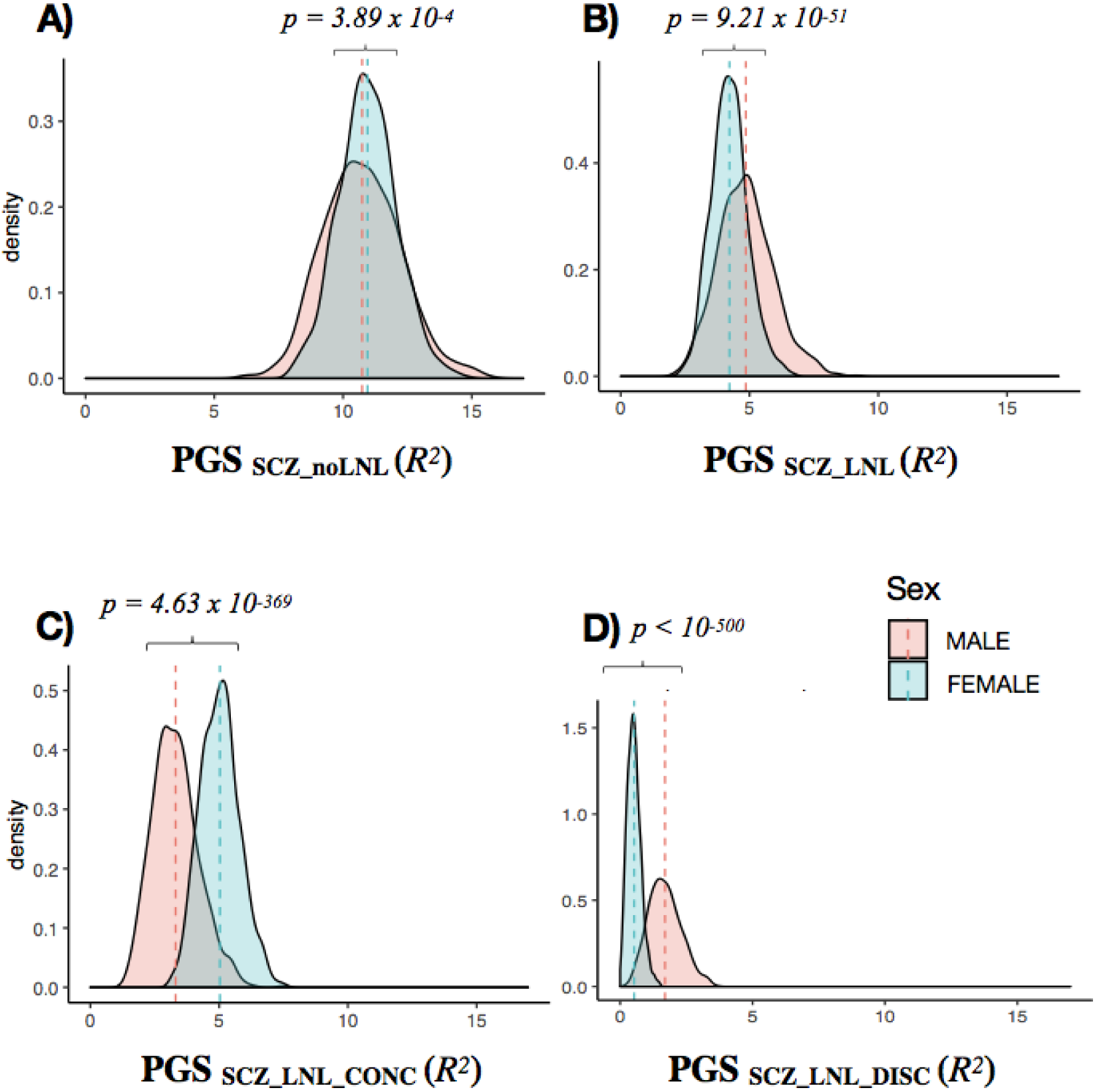
Density plot for sex comparison of PGS_SCZ_ predictions. 5000 permutations of PGS_SCZ_ predictions in case - control subsamples of 500 schizophrenia patients (SCZ) and 500 healthy controls (HC) (selected from the overall CIBERSAM case-control sample) were performed in males (N_SCZ_ = 1253; N_HC_= 859) and females (N_SCZ_=674; N_HC_= 702) separately. Mean SCZ-HC variance explained by PGS_SCZ_ in males and females was compared for predisposing variation within A) **SCZ_noLNL**, B) **SCZ_LNL**, C) **SCZ_LNL_CONC**, and D) **SCZ_LNL_DISC**. Variance explained in females and males was statistically compared with two-sided t-tests.

### Covariance between SCZ and related phenotypes based on LNL-ISO

We assessed covariance between predisposing genetic variation to schizophrenia and a series of neuropsychiatric disorders and related phenotypes across **SCZ_noLNL, SCZ_LNL_CONC**, and **SCZ_LNL_DISC** using GNOVA^45^.

The majority of the disorders (MDD, ANX, ADHD, ASD, CROSS-DIS, ALC-DEP) and personality traits (NEUR, SWB, DS, PSY_EXP) tested here showed genetic positive correlation within **SCZ_LNL_CONC** and genetic negative correlation within **SCZ_LNL_DISC** (**Figure 3**). However, BIP and OCD showed positive covariances within both genomic annotations. As expected, estimated correlations within **SCZ_noLNL** were similar to those previously described for schizophrenia across the whole genome ^55^ **(Figure 3, Supplementary Table 5)**.

**Figure 3.**
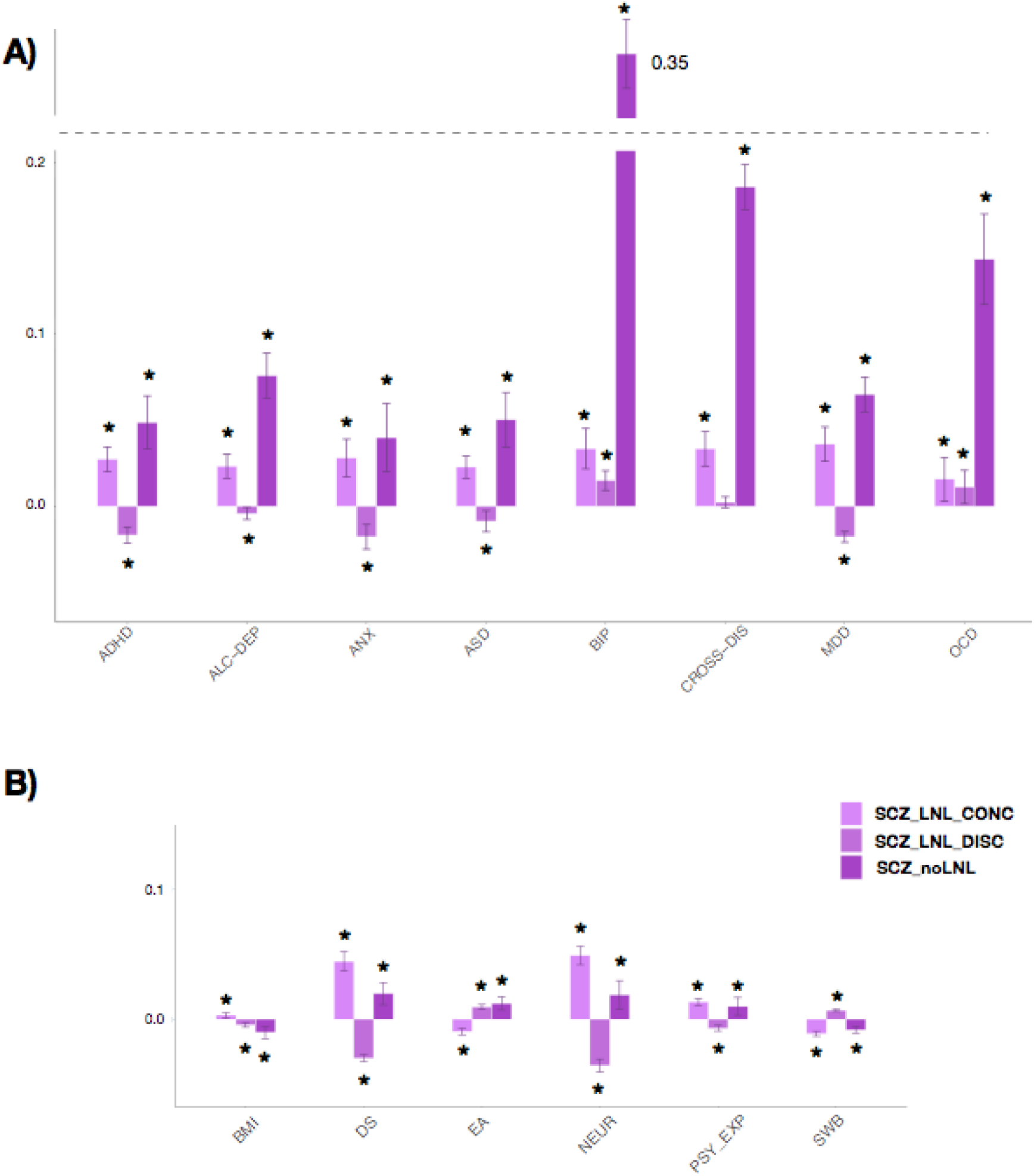
Genetic covariance between schizophrenia and other relevant disorders (A) and related traits (B) across LNL-ISO annotations. Covariances were calculated with GNOVA within SNP subsets from SCZ_noLNL, SCZ_LNL_CONC, and SCZ_LNL_DISC. Error bars represent 95% confidence intervals. FDR-corrected significant associations (*p*_*FDR*_ < 0.05) are marked with an asterisk. Traits and disorders are abbreviated as follows: major depression (MDD), attention and deficit hyperactivity disorder (ADHD), autism spectrum disorder (ASD), anxiety disorder (ANX), Bipolar disorder (BIP), obsessive compulsive disorder (OCD), alcohol dependence disorder (ALC-DEP), cross-disorder phenotype (CROSS-DIS), neuroticism (NEUR), depressive symptoms (DS), subjective well-being (SWB), psychotic experiences in the general population (PSY-EXP), educational attainment (EA) and body-mass index (BMI).

### Mendelian randomization analyses of LNL-ISO and schizophrenia

We found evidence for a strong causal effect of LNL-ISO on schizophrenia (WM - β (standard error (se)) = 1.37 (0.40), p = 6.14 × 10^−4^; CAUSE - γ (CI95%) = 0.61 (0.34, 0.89), p = 0.003) and, to a lesser degree, for a causal effect of schizophrenia liability on LNL-ISO (WM - β (se) = 0.015(0.005), p = 0.008; CAUSE - γ (CI95%) = 0.01 (0.01, 0.01), p = 0.003; table 1). MR-Egger intercept had non-significant, non-zero effects (p = 0.36). We observed evidence of heterogeneity, with some significant SNPs not following the general trend (**Supplementary Figure 4**) which justified the preference for the WM method **(see Supplementary Methods and Table 1)**. Using MR-PRESSO^53^, which eliminates outliers that may be influencing the outcome due to pleiotropy, the effect size of LNL-ISO on schizophrenia and the statistical significance increased **(Table 1)**.

We found comparable evidence for bidirectional causality between Loneliness UKBB and schizophrenia to that found for LNL-ISO (**Table 1, Supplementary Figure 5**). We also found a unidirectional negative causal effect of *ability to confide* on schizophrenia (WM - β (se) = -0.6 (0.19), p = 0.002), and a unidirectional negative causal effect of schizophrenia on *number of people in household* (WM - β (se) = -0.011 (0.003), p = 5.86 × 10^−3^). We found no evidence of causality between the number of family/friends visits and schizophrenia.

## DISCUSSION

This work suggests the presence of genetic overlap between a social isolation and schizophrenia, with a bidirectional causal relationship between the two phenotypes. We found that overlapping predisposing genetic variation with concordant effects in both phenotypes shows significant SNP-based heritability enrichment, thus supporting the relatively enhanced contribution of this set of variants to schizophrenia liability. Evaluating the role of concordant and discordant genetic variation in schizophrenia, we described concordant variation to be more predictive in females and positively correlated with other neuropsychiatric traits. Conversely, discordant variation was only significantly predictive in males and negatively correlated with most neuropsychiatric traits. Exceptionally, OCD and BIP were positively correlated with schizophrenia regardless of the LNL-ISO-based annotations. These results reveal the likely genomic footprint of social isolation on the heritability of schizophrenia, and provide new insights about their relationship ^33,34^. They also support the role of LNL-ISO as a critical social trait to understand the heterogeneity of pleiotropic genetic effects between schizophrenia and other psychiatric disorders and behavioral traits.

To our knowledge, this is the first study to report a significant polygenic score prediction of social isolation in a schizophrenia sample. We also found a significant polygenic score prediction for each of the individual traits included in the composite LNL-ISO phenotype on schizophrenia. These results mirror the finding of a clinical overlap between schizophrenia and both perceived loneliness and objective social disconnection and support that social isolation may play a significant role in the etiology of psychotic disorders ^17,20,27,56^.

Both polygenic score predictions and LD-score-based partition heritability estimates have been previously described as powerful methods to evaluate the effects of genetic predisposing variation within specific subsets of variants ^43,57,58^. With 3.8% of the SNPs explaining an estimated 13.1% of the SNP-based heritability, concordant overlapping variation between both phenotypes exhibits more than a three-fold increase in heritability enrichment compared to variants not predisposing to LNL-ISO, and a much higher enrichment than most of the genome-wide annotations previously evaluated in schizophrenia ^59^.

Despite reported sex differences in the epidemiology and clinical manifestations of psychotic disorders ^35,37,60^, studies have not found an unequivocal effect of sex on genetic associations so far ^61^. By analyzing the genomic overlap between schizophrenia and LNL-ISO, we did observe a differential effect of sex on polygenic predictions. Concordant overlapping variants in SCZ and LNL-ISO accounted for a significantly greater amount of variance in schizophrenia risk in females, while discordant variation was predictive only in males in our target sample. These results are in line with recent studies suggesting a potentially higher impact of loneliness and objective social isolation on psychiatric outcomes in females than in males ^26,40^. Moreover, among patients with schizophrenia, loneliness has been described to be more prevalent in females than males ^62^.

Genetic correlations have been shown to be a very useful method for understanding shared genetic architecture and the interrelationship between disorders and related traits, even though it still maintains certain limitations ^42,55,63,64^. By evaluating local genetic correlations, previous studies have described subtle structures in shared genetics between complex traits ^45,65^. Even in schizophrenia or ASD some counterintuitive correlations with related disorders arise within specific subsets of variants ^65,66^. In our study, we have also described the impact of the genetic liability to LNL-ISO in the relationship between schizophrenia and most of the tested neuropsychiatric disorders (ASD, MDD, ANX, ADHD, ALC-DEP) and other related behavioural traits (SWB, NEUR, PSY-EXP, EA). In the majority of the disorders, schizophrenia is positively correlated within concordant overlapping variation and negatively correlated within discordant overlapping variation with LNL-ISO, thus pointing to a shared genetic impact of social isolation on comorbidity with these disorders. However, OCD and BIP are positively correlated with schizophrenia regardless of LNL-ISO based annotations, thus suggesting that these disorders are independently affected by the genetic predisposition to LNL-ISO. These results are in line with recent findings suggesting that schizophrenia, BIP, and OCD could belong to the same psychopathology factor at the genomic level ^67^.

The genetic relationship between schizophrenia and EA and other cognitive-related measures such as intelligence has been widely studied ^68,69,66^. Assessing partial covariances between EA and schizophrenia, we described a negative covariance within concordant overlapping variation, while EA was positively correlated with schizophrenia across discordant overlapping variation and with variants only associated with schizophrenia (SCZ_noLNL). Our findings suggest that poor educational attainment often found in patients with schizophrenia during childhood and adolescence ^70,71^ could be mediated by a social isolation component.

Mendelian randomization analyses provided evidence of the bidirectional nature of the causal relationship between loneliness and isolation and schizophrenia liability, with a greater size of the effect of LNL-ISO on schizophrenia risk than in the opposite direction. This finding of bidirectional causality between social isolation and schizophrenia was confirmed with the recently developed method CAUSE which provides better control for correlated and uncorrelated horizontal pleiotropy ^54^. Our results are consistent with previous evidence suggesting that loneliness and objective social isolation could trigger both positive and negative psychotic symptoms in clinical and non-clinical populations^17,27^. It could also explain the high levels of loneliness and isolation in individuals at ultra-high risk (UHR) for psychosis, which are present before onset of the established illness ^72^. On the other hand, the described effect of schizophrenia liability on social isolation could also give an explanation to the high prevalence of loneliness in the chronic stages of psychotic illnesses ^17,20,26^.

Causal inferences assessing the relationships between LNL-ISO constituents and schizophrenia also found a unidirectional negative causal *effect of ability to confide* on schizophrenia, in line with recent studies describing the association of lack of confidence and loneliness with psychosis, which may be mediated by negative schemata of others ^29,73^. Moreover, an unidirectional negative causal effect of schizophrenia liability on the *number of people living in your household* was found, thus suggesting a possible indirect causal effect of schizophrenia genetic liability on subsequent social disconnection in participants diagnosed with schizophrenia^74^.

Our study was subject to several limitations. First, we used measures of loneliness and objective social isolation from the UKBB, which are based on single-question questionnaires and not on validated scales such as UCLA Loneliness ^75^. Second, since we used discovery samples for polygenic score predictions from UKBB, population or socio-economic biases could have affected our genetic predictions to some extent ^76,77^. Third, partitioning the genome in order to estimate heritability enrichment in a reduced subset of SNP may have underpowered some of our analyses, thus calling for larger sample sizes in future studies addressing this question. Fourth, we found a great degree of heterogeneity in the MR analyses. However, we implemented several complementary methods to support robustness of our findings and report only on results found to be consistent across methods.

In summary, our results shed additional light on the relationship between social isolation and schizophrenia from a genetic perspective, and lend further support for the potential role of loneliness and social isolation in the onset and maintenance of schizophrenia and other psychotic disorders ^78^. We also provide new insights into the influence of social isolation on comorbidity with other mental disorders and its interplay with behavioral traits. It should be noted that the recent COVID-19 pandemic has heightened levels of loneliness and social isolation across the entire human population, which may likely result in an increase in the risk of developing psychotic and other mental disorders ^79^. Henceforth, considering that social isolation and perceived loneliness may be potentially modifiable, they could become targets for effective preventive interventions with a substantial impact on mental health in the coming years.

## Data Availability

The data that support the findings of this study are openly available in Psychiatric Genomics Consortium website at https://www.med.unc.edu/pgc/download-results/, in Social Science Genetic Association Consortium at https://www.thessgac.org/data and Elucidating the genetic basis of social interaction and isolation at https://doi.org/10.1038/s41467-018-04930-1

## ACKNOWLEDGEMENTS

This work was supported by the Spanish Ministry of Science and Innovation. Instituto de Salud Carlos III (SAM16PE07CP1, PI16/02012, PI19/024), co-financed by ERDF Funds from the European Commission, “A way of making Europe”, CIBERSAM. Madrid Regional Government (B2017/BMD-3740 AGES-CM-2), European Union Structural Funds. European Union Seventh Framework Program under grant agreements FP7-4-HEALTH-2009-2.2.1-2-241909 (Project EU-GEI), FP7-HEALTH-2013-2.2.1-2-603196 (Project PSYSCAN) and FP7-HEALTH-2013-2.2.1-2-602478 (Project METSY); and European Union H2020 Program under the Innovative Medicines Initiative 2 Joint Undertaking (grant agreement No 115916, Project PRISM, and grant agreement No 777394, Project AIMS-2-TRIALS), Fundación Familia Alonso, Fundación Alicia Koplowitz and Fundación Mutua Madrileña. C. M. Díaz-Caneja holds a Juan Rodés Grant from Instituto de Salud Carlos III (JR19/00024). Xaquín Gurriarán has received a grant from Fundación Instituto Roche

## COI

Dr. Arango has been a consultant to or has received honoraria or grants from Acadia, Angelini, Gedeon Richter, Janssen Cilag, Lundbeck, Minerva, Otsuka, Roche, Sage, Servier, Shire, Schering Plough, Sumitomo Dainippon Pharma, Sunovion and Takeda.

Dr. Crespo-Facorro has received honoraria (advisory board and educational lectures) and travel expenses from Takeda, Menarini, Angelini, Teva, Otsuka, Lundbeck and Johnson & Johnson. He has also received unrestricted research grants from Lundbeck.

Dr. Díaz-Caneja has received honoraria from AbbVie, Sanofi, and Exeltis.

## Notes

### Author Declarations

Access to human genomic data will be provided to research investigators who, along with their institutions, have certified their agreement with the expectations and terms of access detailed below. NIH expects that, through Data Access Request (DAR) process, approved users of controlled-access datasets recognize any restrictions on data use established by the Submitting Institutions through the Institutional Certification, and as stated on the dbGaP study page.

## REFERENCES

1. Hughes, M. E., Waite, L. J., Hawkley, L. C. & Cacioppo, J. T. A Short Scale for Measuring Loneliness in Large Surveys: Results From Two Population-Based Studies. Res Aging 26, 655–672 (2004).

2. Shankar, A., Hamer, M., McMunn, A. & Steptoe, A. Social isolation and loneliness: relationships with cognitive function during 4 years of follow-up in the English Longitudinal Study of Ageing. Psychosom Med 75, 161–170 (2013).

3. Baumeister, R. F. & Leary, M. R. The need to belong: desire for interpersonal attachments as a fundamental human motivation. Psychol Bull 117, 497–529 (1995).

4. Perissinotto, C. M. & Covinsky, K. E. Living Alone, Socially Isolated or Lonely— What are We Measuring? J Gen Intern Med 29, 1429–1431 (2014).

5. Cacioppo, J. T., Hawkley, L. C., Norman, G. J. & Berntson, G. G. Social isolation. Ann. N. Y. Acad. Sci. 1231, 17–22 (2011).

6. Holt-Lunstad, J., Smith, T. B., Baker, M., Harris, T. & Stephenson, D. Loneliness and Social Isolation as Risk Factors for Mortality: A Meta-Analytic Review. Perspect Psychol Sci 10, 227–237 (2015).

7. Cacioppo, J. T., Cacioppo, S., Capitanio, J. P. & Cole, S. W. The neuroendocrinology of social isolation. Annu Rev Psychol 66, 733–767 (2015).

8. Steptoe, A., Shankar, A., Demakakos, P. & Wardle, J. Social isolation, loneliness, and all-cause mortality in older men and women. PNAS 110, 5797–5801 (2013).

9. Beutel, M. E. et al. Loneliness in the general population: prevalence, determinants and relations to mental health. BMC Psychiatry 17, 97 (2017).

10. Hawkley, L. C. & Cacioppo, J. T. Loneliness Matters: A Theoretical and Empirical Review of Consequences and Mechanisms. ann. behav. med. 40, 218–227 (2010).

11. Heinrich, L. M. & Gullone, E. The clinical significance of loneliness: a literature review. Clin Psychol Rev 26, 695–718 (2006).

12. Matthews, T. et al. Lonely young adults in modern Britain: findings from an epidemiological cohort study. Psychol Med 49, 268–277 (2019).

13. Meltzer, H. et al. Feelings of loneliness among adults with mental disorder. Soc Psychiatry Psychiatr Epidemiol 48, 5–13 (2013).

14. Peerenboom, L., Collard, R. M., Naarding, P. & Comijs, H. C. The association between depression and emotional and social loneliness in older persons and the influence of social support, cognitive functioning and personality: A cross-sectional study. J Affect Disord 182, 26–31 (2015).

15. Adams, K. B., Sanders, S. & Auth, E. A. Loneliness and depression in independent living retirement communities: risk and resilience factors. Aging Ment Health 8, 475–485 (2004).

16. Cacioppo, J. T., Hughes, M. E., Waite, L. J., Hawkley, L. C. & Thisted, R. A. Loneliness as a specific risk factor for depressive symptoms: cross-sectional and longitudinal analyses. Psychol Aging 21, 140–151 (2006).

17. Michalska da Rocha, B., Rhodes, S., Vasilopoulou, E. & Hutton, P. Loneliness in Psychosis: A Meta-analytical Review. Schizophrenia Bulletin 44, 114–125 (2018).

18. Green, M. F. et al. Social Disconnection in Schizophrenia and the General Community. Schizophr Bull 44, 242–249 (2018).

19. Lim, J. S., Lim, M. Y., Choi, Y. & Ko, G. Modeling environmental risk factors of autism in mice induces IBD-related gut microbial dysbiosis and hyperserotonemia. Molecular Brain 10, (2017).

20. Eglit, G. M. L., Palmer, B. W., Martin, A. S., Tu, X. & Jeste, D. V. Loneliness in schizophrenia: Construct clarification, measurement, and clinical relevance. PLOS ONE 13, e0194021 (2018).

21. van der Werf, M., van Winkel, R., van Boxtel, M. & van Os, J. Evidence that the impact of hearing impairment on psychosis risk is moderated by the level of complexity of the social environment. Schizophr. Res. 122, 193–198 (2010).

22. Bucci, P. et al. Premorbid academic and social functioning in patients with schizophrenia and its associations with negative symptoms and cognition. Acta Psychiatr Scand 138, 253–266 (2018).

23. Kraepelin, E., Robertson, G. M. & Barclay, R. M. Dementia praecox and paraphrenia. (Chicago Medical Book Co., 1919).

24. Mishlove, M. & Chapman, L. J. Social anhedonia in the prediction of psychosis proneness. Journal of Abnormal Psychology 94, 384–396 (1985).

25. Conrad, K. Die beginnende Schizophrenie; Versuch einer Gestaltanalyse des Wahns. (Thieme, 1958).

26. Solmi, M. et al. Factors Associated With Loneliness: An Umbrella Review Of Observational Studies. Journal of Affective Disorders 271, 131–138 (2020).

27. Chau, A. K. C., Zhu, C. & So, S. H.-W. Loneliness and the psychosis continuum: a meta-analysis on positive psychotic experiences and a meta-analysis on negative psychotic experiences. Int Rev Psychiatry 31, 471–490 (2019).

28. Gollwitzer, A., Wilczynska, M. & Jaya, E. S. Targeting the link between loneliness and paranoia via an interventionist-causal model framework. Psychiatry Research 263, 101–107 (2018).

29. Lamster, F., Lincoln, T. M., Nittel, C. M., Rief, W. & Mehl, S. The lonely road to paranoia. A path-analytic investigation of loneliness and paranoia. Comprehensive Psychiatry 74, 35–43 (2017).

30. Abdellaoui, A. et al. Phenome-wide investigation of health outcomes associated with genetic predisposition to loneliness. Hum. Mol. Genet. 28, 3853–3865 (2019).

31. Matthews, T. et al. Social isolation, loneliness and depression in young adulthood: a behavioural genetic analysis. Soc Psychiatry Psychiatr Epidemiol 51, 339–348 (2016).

32. Turley, P. et al. Multi-trait analysis of genome-wide association summary statistics using MTAG. Nat. Genet. 50, 229–237 (2018).

33. Day, F. R., Ong, K. K. & Perry, J. R. B. Elucidating the genetic basis of social interaction and isolation. Nat Commun 9, 2457 (2018).

34. Abdellaoui, A. et al. Predicting loneliness with polygenic scores of social, psychological and psychiatric traits. Genes, Brain and Behavior 17, e12472 (2018).

35. Riecher-Rössler, A., Butler, S. & Kulkarni, J. Sex and gender differences in schizophrenic psychoses—a critical review. Archives of Women’s Mental Health (2018) doi:10.1007/s00737-018-0847-9.

36. Szymanski, S. et al. Gender differences in onset of illness, treatment response, course, and biologic indexes in first-episode schizophrenic patients. Am J Psychiatry 152, 698–703 (1995).

37. Ochoa, S., Usall, J., Cobo, J., Labad, X. & Kulkarni, J. Gender Differences in Schizophrenia and First-Episode Psychosis: A Comprehensive Literature Review. Schizophr Res Treatment 2012, (2012).

38. Barreto, M. et al. Loneliness around the world: Age, gender, and cultural differences in loneliness. Personality and Individual Differences 110066 (2020) doi:10.1016/j.paid.2020.110066.

39. Galderisi, S., Bucci, P., Üçok, A. & Peuskens, J. No gender differences in social outcome in patients suffering from schizophrenia. European Psychiatry 27, 406–408 (2012).

40. Liu, H., Zhang, M., Yang, Q. & Yu, B. Gender differences in the influence of social isolation and loneliness on depressive symptoms in college students: a longitudinal study. Soc Psychiatry Psychiatr Epidemiol 55, 251–257 (2020).

41. Schizophrenia Working Group of the Psychiatric Genomics Consortium. Biological insights from 108 schizophrenia-associated genetic loci. Nature 511, 421–427 (2014).

42. Bulik-Sullivan, B. K. et al. LD Score regression distinguishes confounding from polygenicity in genome-wide association studies. Nature Genetics 47, 291–295 (2015).

43. Finucane, H. K. et al. Heritability enrichment of specifically expressed genes identifies disease-relevant tissues and cell types. Nat Genet 50, 621–629 (2018).

44. Cahoy, J. D. et al. A Transcriptome Database for Astrocytes, Neurons, and Oligodendrocytes: A New Resource for Understanding Brain Development and Function. J. Neurosci. 28, 264–278 (2008).

45. Lu, Q. et al. A Powerful Approach to Estimating Annotation-Stratified Genetic Covariance via GWAS Summary Statistics. Am J Hum Genet 101, 939–964 (2017).

46. Ruth Mitchell, E. MRC IEU UK Biobank GWAS pipeline version 2. data.bris https://data.bris.ac.uk/data/dataset/pnoat8cxo0u52p6ynfaekeigi (2019) doi:10.5523/bris.pnoat8cxo0u52p6ynfaekeigi.

47. Davey Smith, G. & Hemani, G. Mendelian randomization: genetic anchors for causal inference in epidemiological studies. Hum. Mol. Genet. 23, R89–98 (2014).

48. Burgess, S., Dudbridge, F. & Thompson, S. G. Combining information on multiple instrumental variables in Mendelian randomization: comparison of allele score and summarized data methods. Stat Med 35, 1880–1906 (2016).

49. Bowden, J., Davey Smith, G., Haycock, P. C. & Burgess, S. Consistent Estimation in Mendelian Randomization with Some Invalid Instruments Using a Weighted Median Estimator. Genet. Epidemiol. 40, 304–314 (2016).

50. Bowden, J., Davey Smith, G. & Burgess, S. Mendelian randomization with invalid instruments: effect estimation and bias detection through Egger regression. Int J Epidemiol 44, 512–525 (2015).

51. Hartwig, F. P., Davey Smith, G. & Bowden, J. Robust inference in summary data Mendelian randomization via the zero modal pleiotropy assumption. Int J Epidemiol 46, 1985–1998 (2017).

52. Hemani, G. et al. The MR-Base platform supports systematic causal inference across the human phenome. eLife 7, e34408 (2018).

53. Verbanck, M., Chen, C.-Y., Neale, B. & Do, R. Detection of widespread horizontal pleiotropy in causal relationships inferred from Mendelian randomization between complex traits and diseases. Nat. Genet. 50, 693–698 (2018).

54. Morrison, J., Knoblauch, N., Marcus, J. H., Stephens, M. & He, X. Mendelian randomization accounting for correlated and uncorrelated pleiotropic effects using genome-wide summary statistics. Nat. Genet. 52, 740–747 (2020).

55. Bulik-Sullivan, B. et al. An atlas of genetic correlations across human diseases and traits. Nat. Genet. 47, 1236–1241 (2015).

56. Lim, M. H., Gleeson, J. F. M., Alvarez-Jimenez, M. & Penn, D. L. Loneliness in psychosis: a systematic review. Soc Psychiatry Psychiatr Epidemiol 53, 221–238 (2018).

57. Wray, N. R. et al. Research review: Polygenic methods and their application to psychiatric traits. J Child Psychol Psychiatry 55, 1068–1087 (2014).

58. Rammos, A., Gonzalez, L. A. N., Weinberger, D. R., Mitchell, K. J. & Nicodemus, K. K. The role of polygenic risk score gene-set analysis in the context of the omnigenic model of schizophrenia. Neuropsychopharmacology 44, 1562–1569 (2019).

59. Finucane, H. K. et al. Partitioning heritability by functional annotation using genome-wide association summary statistics. Nat. Genet. 47, 1228–1235 (2015).

60. Häfner, H. et al. The Epidemiology of Early Schizophrenia: Influence of Age and Gender on Onset and Early Course. British Journal of Psychiatry 164, 29–38 (1994).

61. Consortium, S. W. G. of the P. G., Ripke, S., Walters, J. T. & O’Donovan, M. C. Mapping genomic loci prioritises genes and implicates synaptic biology in schizophrenia. medRxiv 2020.09.12.20192922 (2020) doi:10.1101/2020.09.12.20192922.

62. Badcock, J. C. et al. Loneliness in psychotic disorders and its association with cognitive function and symptom profile. Schizophrenia Research 169, 268–273 (2015).

63. Lee, J. J., McGue, M., Iacono, W. G. & Chow, C. C. The accuracy of LD Score regression as an estimator of confounding and genetic correlations in genome-wide association studies. Genet Epidemiol 42, 783–795 (2018).

64. Ni, G., Moser, G., Wray, N. R. & Lee, S. H. Estimation of Genetic Correlation via Linkage Disequilibrium Score Regression and Genomic Restricted Maximum Likelihood. Am J Hum Genet 102, 1185–1194 (2018).

65. Zhang, Y. et al. Local genetic correlation analysis reveals heterogeneous etiologic sharing of complex traits. bioRxiv 2020.05.08.084475 (2020) doi:10.1101/2020.05.08.084475.

66. Lam, M. et al. Pleiotropic Meta-Analysis of Cognition, Education, and Schizophrenia Differentiates Roles of Early Neurodevelopmental and Adult Synaptic Pathways. The American Journal of Human Genetics 105, 334–350 (2019).

67. Waldman, I. D., Poore, H. E., Luningham, J. M. & Yang, J. Testing structural models of psychopathology at the genomic level. World Psychiatry 19, 350–359 (2020).

68. Escott-Price, V. et al. Genetic liability to schizophrenia is negatively associated with educational attainment in UK Biobank. Mol. Psychiatry 25, 703–705 (2020).

69. Le Hellard, S. et al. Identification of Gene Loci That Overlap Between Schizophrenia and Educational Attainment. Schizophr Bull 43, 654–664 (2017).

70. Hakulinen, C. et al. The association between early-onset schizophrenia with employment, income, education, and cohabitation status: nationwide study with 35 years of follow-up. Soc Psychiatry Psychiatr Epidemiol 54, 1343–1351 (2019).

71. Dickson, H. et al. Academic achievement and schizophrenia: a systematic meta-analysis. Psychological Medicine 50, 1949–1965 (2020).

72. Robustelli, B. L., Newberry, R. E., Whisman, M. A. & Mittal, V. A. Social relationships in young adults at ultra high risk for psychosis. Psychiatry Res 247, 345–351 (2017).

73. Müller, H. et al. Negative schemata about the self and others and paranoid ideation in at-risk states and those with persisting positive symptoms. Early Interv Psychiatry 12, 1157–1165 (2018).

74. Green, M. F. et al. Social Disconnection in Schizophrenia and the General Community. Schizophrenia Bulletin 44, 242–249 (2018).

75. Russell, D. W. UCLA Loneliness Scale (Version 3): Reliability, Validity, and Factor Structure. Journal of Personality Assessment 66, 20–40 (1996).

76. Fry, A. et al. Comparison of Sociodemographic and Health-Related Characteristics of UK Biobank Participants With Those of the General Population. Am. J. Epidemiol. 186, 1026–1034 (2017).

77. Haworth, S. et al. Apparent latent structure within the UK Biobank sample has implications for epidemiological analysis. Nature Communications 10, 333 (2019).

78. Bennett, J. C., Surkan, P. J., Moulton, L. H., Fombonne, E. & Melchior, M. Childhood social isolation and psychotic experiences in young adulthood: a community based study. Eur Child Adolesc Psychiatry 29, 1003–1010 (2020).

79. Moreno, C. et al. How mental health care should change as a consequence of the COVID-19 pandemic. The Lancet Psychiatry 7, 813–824 (2020).

