## Supplementary Methods for "Unraveling the relationship of loneliness and isolation in schizophrenia: Polygenic dissection and causal inference"

**Supplementary Methods 4. LD-score regression (LDSC) and partitioning SNP heritability ……………………………………………………………………........................................................ 6**

**Supplementary Methods 5. Partial genetic correlations between SCZ and related traits based on LNL-ISO annotations ……………………………………………………………………………….. 8**

**Supplementary Methods 6. Bidirectional Causal Analyses: Two-sample Mendelian Randomization ……..………………………………………………………………………………….. 9**

**Supplementary Methods 1**. **Participants and QC filtering**

Blood samples were collected from 2145 subjects with a diagnosis of DSM-IV schizophrenia spectrum disorders or schizoaffective disorder (SCZ) and 1711 healthy controls (HC) at 8 different Hospitals across Spain that belong to CIBERSAM (Spain, https://www.cibersam.es/en). Individual genetic data was obtained by genotyping as part of the third wave of the Schizophrenia genome wide association study (GWAS) performed by the Psychiatric Genomics Consortium (PGC).

After QC filtering, genetic data from 1927 SCZ (65% males) and 1561 HC (55% males) were used for the subsequent analyses. All remaining subjects were of European Ancestry. Average age at inclusion was 33.15 years old.

### Quality Control and imputation

QC filters were applied following PGC-SZ2 criteria^1^. Only autosomal genetic data was used. Briefly, consecutive filters were applied as follows:

- SNP missingness across samples < 0.05 (Before sample removal)

- Removal of subjects with a SNP missingness > 0.02.

- Autosomal heterozygosity deviation (|F het | < 0.2)

- SNP missingness < 0.02 (after sample removal)

- Difference in SNP missingness between SZ and HC < 0.02

- Hardy-Weinberg (HW) equilibrium (P > 10^-6^ in HC or P > 10^-10^ in SZ)

Michigan imputation server (https://imputationserver.sph.umich.edu/index.html) was used for imputation in order to gain genetic information beyond genotyped variants. Finally, 9340561 variants with imputation quality > 0.3 and MAF > 0.01 and 3489 individuals (1927 SZ and 1561 HC) were retained.

Multidimensional scaling (MDS) was used to generate ancestry MDS covariates after imputation using variants with high imputation quality (INFO >0.8) , MAF > 0.01 and after removal of Major histocompatibility complex variation (from 26Mb to 33Mb of chromosome 6). SNP were pruned using PLINK v.1.9 with r2 < 0.1 in 500 SNP (*--indep-pairwise 500 1 0.1*). We used the 10 first MDS components as covariates for polygenic scores prediction in this case - control cohort.

**Supplementary Methods 2**. **Summary statistics from loneliness and isolation traits**

Summary statistics for the loneliness and social isolation composite (LNL-ISO) previously explored^2^ were downloaded from the available repository (https://doi.org/10.17863/CAM.23511). The authors performed a combined multi-trait GWAS (MTAG) in the UK Biobank study yielding an effective sample size of 487,647 individuals. Briefly, they combined summary genetic data from three GWAS at the UK Biobank sample, derived from three questions related to loneliness and social isolation:

(a) 'Do you often feel lonely?', as if individuals answered 'yes' (recorded as cases) or 'no' (controls)

(b) A composite variable based on the questions 'Including yourself, how many people are living together in your household?' and ''How often do you visit friends or family or have them visit you?' (cases were defined as those who lived alone and who indicated that they either never visited or had no friends or family outside their household; controls were defined as those who either did not live alone or had friends who visited at least once a week).

(c) A variable representing the quality of social interactions 'How often are you able to confide in someone close to you?' (cases were defined as those who answered 'Never or almost never', controls were defined as those who answered 'Almost daily').

Summary data from the latest UK biobank (UKBB) results regarding loneliness and isolation traits, from which LNL-ISO MTAG was originally created, were used: Loneliness (https://nealelab.github.io/UKBB_ldsc/h2_summary_2020.html), ability to confide (https://nealelab.github.io/UKBB_ldsc/h2_summary_2110.html) and social support measured by number of family and friend visits (https://nealelab.github.io/UKBB_ldsc/h2_summary_1031.html) and number of people living in household (<https://nealelab.github.io/UKBB_ldsc/h2_summary_709.html>).

**Supplementary Methods 3**. **Polygenic score predictions**

Discovery datasets for Polygenic score calculations were filtered for imputation quality score > 0.8, include only biallelic variation and exclude indels. Correlated SNPs due to linkage disequilibrium (LD) were removed using PLINK 1.9 clumping algorithm (r^2^ > 0.1; window size = 500 kb). Due to the extremely complex LD pattern, genetic variants within Major Histocompatibility Complex (MHC) were removed (from 26Mb to 33Mb of chromosome 6). PLINK 1.9 was used to calculate PGS across schizophrenia patients and healthy controls weighted by the logOR in the discovery sample. Several P thresholds were used (P<5 x 10^-8^, 5 x 10^-5^, 1 x 10^-3^, 0.01, 0.05, 0.1, 0.2, 0.5 and 1).

Polygenic models were performed based on LNL-ISO MTAG^2^ and PGC-SZ2^1^ GWAS summary statistics as discovery sample, and SCZ_CIBERSAM case-control cohort was used as target sample. Standardised PGS were calculated and significance was evaluated by logistic regression, using case-control status as dependent variable and sex, age and 10 first MDS ancestry components as covariates. Explained variance attributable to PGS was calculated as the increase in Naggelkerke’s pseudo-R^2^ between a model with and without PGS variable. All p-values were FDR corrected by Benjamini - Hochberg method (*p_FDR_*) and the most significant P-threshold was confirmed after 10K permutations of the case control status. The variance explained in the real case (estimated as Naggelkerke’s pseudo-R^2^) was compared to the distribution of 10K generated explained variances under randomly generated scenarios based on random permutations of the case-control status. P value after permutations (*p_perm_*) was estimated as the probability to obtain, across the generated distribution, a higher percentage of explained variance than in the real case.

In order to analyze the role of PGS_SCZ_ in the prediction of SCZ - HC status but taking into account the role of the genetic variation in LNL-ISO, SCZ summary data was divided into three different subsets of variants, according to their role in LNL-ISO. First, only variants included both in SCZ and LNL-ISO summary data were included (Final N_SNPs = 5658282). Second, predisposing variation to SCZ was divided into variants not associated with LNL-ISO (**SCZ_noLNL**; P_LNL-ISO_ > 0.05; N_SNPs = 5172017) and variants associated with LNL-ISO (**SCZ_LNL**; P_LNL-ISO_ < 0.05; N_SNPs = 486265). Third, SCZ_LNL was again divided into those variants with concordant (**SCZ_LNL_CONC;** N_SNPs = 269361) and discordant (**SCZ_LNL_DISC;** N_SNPs = 216904) sign of the effect allele between SCZ and LNL-ISO. In each dataset, correlated SNPs due to linkage disequilibrium (LD) were removed using PLINK 1.9 clumping algorithm described before and independent variants within **SCZ_noLNL** (N_clumpedSNPs = 169574), **SCZ_LNL** (N_clumpedSNPs = 11804), **SCZ_LNL_CONC** (N_clumpedSNPs = 6468) and **SCZ_LNL_DISC** (N_clumpedSNPs = 5336) were used to calculate PGS on the SCZ_CIBERSAM case-control cohort (N_SCZ = 1927; N_HC = 1561). Standardised PGS were calculated and significance was evaluated by logistic regression, using case-control status as dependent variable and sex, age and 10 first MDS ancestry components as covariates. Explained variance attributable to PGS was calculated as the increase in Naggelkerke’s pseudo-R^2^ between a model with and without PGS variable. All p-values were FDR corrected and the most significant P-threshold was confirmed after 10K permutations of the case control status.

Since loneliness perception and sociability have been described to depend on sex (for instance, higher percentage of cases with loneliness autoperception in UKBB cohort are women(26.5% of women) instead of men(17.3% of men), we were motivated to analyze the impact of schizophrenia predisposing variation in the SCZ_CIBERSAM case-control cohort across the different subset of SNPs based on their role in LNL-ISO. For this purpose, A permutation-based approach was performed: 5000 random selections of 500 SCZ and 500 HC subjects across men and women separately within the SCZ_CIBERSAM cohort were done, and PGS calculation and explained variance predictions were calculated by logistic regressión and Naggelkerke’s pseudo-R^2^ estimations. The process was repeated for each of the aforedescribed SNP subsets: SCZ_noLNL, SCZ_LNL, SCZ_LNL_CONC and SCZ_LNL_DISC. Variance explained in women and men was statistically compared by student t-test.

**Supplementary Methods 4. LD-score regression (LDSC) and partitioning SNP heritability**

To obtain SNP heritability estimates related to annotations studied here, recommended procedure was followed^3,4^ (<https://github.com/bulik/ldsc/wiki/Partitioned-Heritability>).

First, per SNP annotation files were created (one per chromosome and desired annotation). Each file consisted of a row per SNP and a column for each sub-annotation (1 = a SNP is part of that sub-annotation). SNP that do not belong to the annotation were given a value of 0. Annotation files were created for:

- A) Each genome partition based on the relationship between SCZ and LNL-ISO, as done with polygenic score predictions: SCZ_noLNL and SCZ_LNL_CONC. SCZ_LNL_DISC (see supplementary methods 3).
- B) Adapted sub-annotation files for the intersection between the above described annotations (SCZ_noLNL and SCZ_LNL_CONC) and 10 whole tissue^4,5^ , 13 brain-related tissue (Brain GTEx from Finucane et al., 2018^3^) and 3 brain cell-type annotation files (Neuron, astrocytes and oligodendrocytes)^3,6^ available at LDSC repository (<http://data.broadinstitute.org/alkesgroup/LDSCORE/>). SCZ_LNL_DISC was not included here as the percentage of SNP-based heritability explained by this annotation was too low to assess enrichment within sub-annotations generated from it.

In order to generate annotation files from A, bed files were generated from the summary statistics overlap between SCZ and LNL-ISO as described before. In the case of annotation files from B), bed files were initially generated by intersection between bed files from SCZ / LNL- ISO and bed-files from cell-type and tissue available annotations at LDSC repository. Bed file intersection was performed with Bedtools^7^ , using *--intersectBed* command. Once bed files were generated, annotation files were obtained using *--make-annot.py* and data files from phase 3 of the 1000 Genomes Project (1000 Genomes Project Consortium, 2015). A separated file was obtained for each annotation and chromosome.

LDSC was run using associated data files from phase 3 of the 1000 Genomes Project^8^ . LD scores were computed for each annotation file using the recommended parameters: 1-cM window (*--ld-wind-cm* 1), restriction to Hapmap3 SNPs and exclusion of major histocompatibility complex (MHC) region due to its high gene density and exceptional LD , as recommended by the developers^3^ . The ‘*–overlap-annot*’ argument and 1000 genomes phase 3 - based frequency files (‘1000G_Phase3_frq’ files via *--frqfile-chr* argument) and LD weights (‘weights_hm3_no_hla’ files via *--w-ld-chr* argument) were used for LD score calculations.

SCZ summary data from previous PGC GWAS^1^ was used as input for heritability enrichment calculation. *--munge-sumstats.py* command was used for formatting summary data, and only SNPs present in HapMap 3 were included.

Partitioned LDSC computes the proportion of SNP heritability associated with each annotation column while taking into account all other annotations. This is performed by regression models using the estimated LD-scores jointly with other independent LD scores for baseline annotations to improve the model performance. The full baseline model v2.2, consistent of a full annotation column (1 per all SNPs) and 158 independent functional annotations, available at LDSC repository (<https://data.broadinstitute.org/alkesgroup/LDSCORE/>), was used as independent LD scores. Indeed, for the heritability estimation for annotations from B), based on intersection between SCZ/ LNL-ISO and tissue/cell-types, an additional independent LD-scores were used, as described next. As discussed with LDSC developers, the best way to obtain heritability estimates for a cell-type subannotation (for instance, neuronal cell-type annotation within SCZ_LNL_CONC) the inclusion of the annotation from which the cell-type is sub-annotated is recommended (in the example case, the SCZ_LNL_CONC annotations), apart from the independent annotations in the full baseline model.

Based on the proportion of total SNPs in an annotation and the percentage of the SNP heritability (*h^2^_SNP_*) in every case, LDSC calculates an enrichment score and an associated enrichment P value. For annotation files in B), heritability was also estimated for the intersection of the provided control files (these are annotations corresponding to all genes included in the study from which specifically expressed cell types are described) and the corresponding SCZ / LNL-ISO annotations. In the case of specific cell-types^6^ and brain expressed genes from GTEx, ‘anti-target cell-type’ enrichments were also computed. These are calculated as the intersection between the cell-type or brain specific annotation and the variation not belonging to the corresponding SCZ / LNL-ISO annotation. By doing this, specific cell-type enrichment within a particular SCZ / LNL - ISO annotation (for instance, neuronal enrichment within SCZ_LNL_CONC annotation) is compared to the cell-type enrichment out of this SCZ / LNL - ISO annotation (for the later case, neuronal enrichment out of SCZ / LNL-ISO annotation), and the influence of LNL-ISO on a particular cell-type enrichment in SCZ could be estimated.

Moreover, LD score regression applied to specifically expressed genes (LDSC-SEG) was also run using *--h2-cts* argument to perform a one-sided t-test for evaluating whether the cell-type enrichment within a particular LNL-ISO annotation is higher than the associated ‘anti-target cell-type’ enrichment described before.

**Supplementary Methods 5. Partial genetic correlations between SCZ and related traits based on LNL-ISO annotations.**

To examine how the LNL-ISO based annotations (SCZ_noLNL, SCZ_LNL_CONC and SCZ_LNL_DISC) influence the correlation between schizophrenia and other related traits or disorder, correlations were performed but restricted to SNP subsets within each annotation. We selected a series of neuropsychiatric disorders and traits that have been previously demonstrated to be significantly correlated with SCZ and/or social isolation phenotypes. Available summary GWAS data from recent performed studies on the following disorders was obtained: major depression (MDD)^9^, attention and deficit hyperactivity disorder (ADHD)^10,11^, autism spectrum disorders (ASD)^12^, Anxiety disorder (ANX)^13^, Bipolar disorder (BIP)^14^, obsessive compulsive disorder (OCD)^15^, alcohol dependence disorder (ALC-DEP)^16^ and cross-disorder (CROSS-DIS)^17^. Also, related traits with available genetic data were also studied: neuroticism (NEUR)^18^, depressive symptoms (DS)^19^, subjective well-being (SWB)^19^, psychotic experiences in the general population (PSY-EXP)^20^, educational attainment (EA)^21^ and body-mass index (BMI)^22^.

Partial correlations were studied with GNOVA^23^. This program implements an approach similar to LD score regression but capable of working with SNP subsets, using LD-score generated files in LDSC for heritability estimation again in this step. GNOVA calculates the covariance between two disorders with a procedure that accounts for the LD structure and that is demonstrated to be robust to sample overlap (<https://github.com/xtonyjiang/GNOVA>). Full covariance between SCZ and the analysed traits here independent of annotations was also performed with LD score regression program (by the *--rg* argument). Similar to LDSC procedure, 1000 genomes phase 3 and hapmap3 files were used as reference files. *--munge-sumstats.py* program from LDSC was used to generate the formatted summary statistics to be used as input files for GNOVA. As recommended by the GNOVA developers, covariance instead of correlation estimates were used. Derived p-values were statistically corrected by Benjamini-Hochberg FDR procedure (*p_FDR_*).

**Supplementary Methods 6. Bidirectional Causal Analyses: Two-sample Mendelian Randomization**

We used Mendelian Randomization to investigate the bidirectional causal relationships between Loneliness and Isolation traits and Schizophrenia liability.

Mendelian randomization (MR) is a statistical method for inferring causal effects that utilizes genetic variants as Instrumental Variables (IV) that are robustly associated with (a potentially modifiable) exposure to another outcome^24^.

Valid Instrumental variables in Two-sample Mendelian randomization relies on three main assumptions: 1) IV must be associated with the exposure (the relevance assumption); 2) IV must be independent of confounders and share no common mediator in the exposure-outcome relationship (the independence assumption); and 3) The IV affects the outcome only through their effects on the exposure (exclusion restriction assumption)^25^.

These assumptions are very restrictive so different methods have been developed to deal with the violation of these assumptions, especially to control horizontal pleiotropy. Horizontal pleiotropy is a major threat to the validity of an MR analysis. For this reason we use different MR methods to deal with possible biases, and we use a recently developed method (CAUSE) for accounting of correlated and uncorrelated pleiotropy^26^

We have used the data of “Loneliness UKBB”, “Frequency of family visits”, “Number of people in household” and “able to confide” from MR-BASE ([https://gwas.mrcieu.ac.uk](https://gwas.mrcieu.ac.uk/)), based on an API wrapper (<https://mrcieu.github.io/ieugwasr/>) integrated in TwosampleMR package ([https://mrcieu.github.io/TwoSampleMR](https://mrcieu.github.io/TwoSampleMR/))^27^, to conduct MR isolation-traits analyses. The latest GWAS analysis of IEU GWAS consortium^28^ were used and summary data have been obtained of GWAS VCF files with the [gwasvcf R package](https://github.com/mrcieu/gwasvcf) (<https://github.com/mrcieu/gwasvcf>)

The five classic MR methods- Inverse Variance Weighted (IVW)^24^, Weighted Median^29^, MR-Eggers^30^, Simple Mode and Weighted mode^31^- were conducted in the R package TwoSampleMR v.0.5.3 (<https://github.com/mrcieu/TwoSampleMR>) with the default settings.

TwoSampleMR used a default LD-Clumping R2 threshold of 0.001 and a window of 10000kb

Presence of pleiotropy was examined by the MR–Egger intercept test, where a significant non-zero intercept (p < 0.05) indicates horizontal pleiotropy. Heterogeneity test and other sensitivity analyses as leave-one-out test and Single SNP test were also implemented (S**upplementary figure 6**).

Additionally, we conducted Mendelian Randomization Pleiotropy RESidual Sum and Outlier (MR-PRESSO)^32^ (<https://github.com/rondolab/MR-PRESSO>) for accounting for horizontal pleiotropy by outliers removal. MR-PRESSO used RADIAL MR test^33^ to identify outliers and allowed them to remove them. We implemented MR-PRESSO with a wrapped function of TwoSampleMR with NbDistribution = 1000 and DistortionTest SignifThreshold = 0.05 parameters.

Finally, we use a new MR method -CAUSE-^26^ that uses all variables rather than only the most strongly associated with the exposure and takes into account both correlated and uncorrelated pleiotropy. CAUSE uses a mixture model for all the variants and the effect estimates and standard errors measured in GWAS of the two traits. Using the same parameters the authors have used to explain the method, if M is the mediator, (exposure) and Y (outcome) where γ is the true causal effect, Zj is an indicator that variant Gj is a correlated pleiotropic and θj is an indicator of uncorrelated pleiotropy, then:
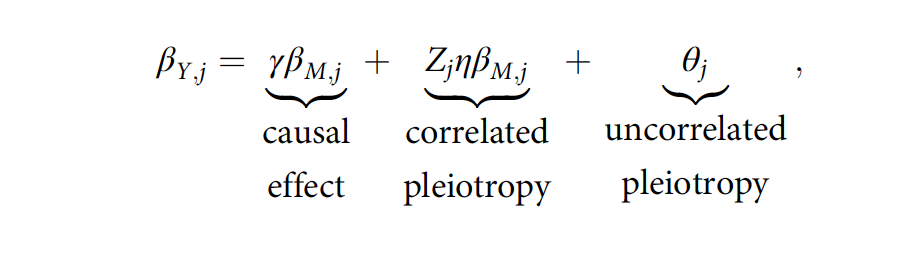

Cause estimated posterior distributions of γ, η and q and compare the fit of posteriors from models with and without a causal effect. For this CAUSE compare a model in which the causal effect is fixed at zero (the sharing model) to a model that allows a nonzero causal effect (the causal model). CAUSE use the expected log pointwise posterior density (ΔELPD), a Bayesian model comparison approach to compare between the two models. We used the recommended default parameters of LD-pruning r2 = 0.1 and the Beta (1,10) prior distribution for q in our analyses (for more details of the method, see (<https://jean997.github.io/cause/>).

In all of our results we have applied multiple testing corrections by Benjamini-Hochberg FDR (P_fdr < 0.05).

**References**

1.	Schizophrenia Working Group of the Psychiatric Genomics Consortium. Biological insights from 108 schizophrenia-associated genetic loci. *Nature* **511**, 421–427 (2014).

2.	Day, F. R., Ong, K. K. & Perry, J. R. B. Elucidating the genetic basis of social interaction and isolation. *Nat Commun* **9**, 2457 (2018).

3.	Finucane, H. K. *et al.* Heritability enrichment of specifically expressed genes identifies disease-relevant tissues and cell types. *Nat Genet* **50**, 621–629 (2018).

4.	Finucane, H. K. *et al.* Partitioning heritability by functional annotation using genome-wide association summary statistics. *Nat. Genet.* **47**, 1228–1235 (2015).

5.	Bulik-Sullivan, B. K. *et al.* LD Score regression distinguishes confounding from polygenicity in genome-wide association studies. *Nature Genetics* **47**, 291–295 (2015).

6.	Cahoy, J. D. *et al.* A Transcriptome Database for Astrocytes, Neurons, and Oligodendrocytes: A New Resource for Understanding Brain Development and Function. *J. Neurosci.* **28**, 264–278 (2008).

7.	Quinlan, A. R. & Hall, I. M. BEDTools: a flexible suite of utilities for comparing genomic features. *Bioinformatics* **26**, 841–842 (2010).

8.	Auton, A. *et al.* A global reference for human genetic variation. *Nature* **526**, 68–74 (2015).

9.	Wray, N. R. *et al.* Genome-wide association analyses identify 44 risk variants and refine the genetic architecture of major depression. *Nature Genetics* **50**, 668–681 (2018).

10.	Demontis, D. *et al.* Discovery of the first genome-wide significant risk loci for attention deficit/hyperactivity disorder. *Nature Genetics* **51**, 63–75 (2019).

11.	Martin, J. *et al.* A Genetic Investigation of Sex Bias in the Prevalence of Attention-Deficit/Hyperactivity Disorder. *Biol Psychiatry* **83**, 1044–1053 (2018).

12.	Grove, J. *et al.* Identification of common genetic risk variants for autism spectrum disorder. *Nat Genet* **51**, 431–444 (2019).

13.	Otowa, T. *et al.* Meta-analysis of genome-wide association studies of anxiety disorders. *Mol Psychiatry* **21**, 1391–1399 (2016).

14.	Stahl, E. A. *et al.* Genome-wide association study identifies 30 loci associated with bipolar disorder. *Nature Genetics* **51**, 793–803 (2019).

15.	Arnold, P. D. *et al.* Revealing the complex genetic architecture of obsessive–compulsive disorder using meta-analysis. *Molecular Psychiatry* **23**, 1181–1188 (2018).

16.	Walters, R. K. *et al.* Transancestral GWAS of alcohol dependence reveals common genetic underpinnings with psychiatric disorders. *Nature Neuroscience* **21**, 1656–1669 (2018).

17.	Cross-Disorder Group of the Psychiatric Genomics Consortium. Electronic address: & Cross-Disorder Group of the Psychiatric Genomics Consortium. Genomic Relationships, Novel Loci, and Pleiotropic Mechanisms across Eight Psychiatric Disorders. *Cell* **179**, 1469-1482.e11 (2019).

18.	Nagel, M. *et al.* Meta-analysis of genome-wide association studies for neuroticism in 449,484 individuals identifies novel genetic loci and pathways. *Nature Genetics* **50**, 920–927 (2018).

19.	Okbay, A. *et al.* Genetic variants associated with subjective well-being, depressive symptoms, and neuroticism identified through genome-wide analyses. *Nature Genetics* **48**, 624–633 (2016).

20.	Legge, S. E. *et al.* Association of Genetic Liability to Psychotic Experiences With Neuropsychotic Disorders and Traits. *JAMA Psychiatry* **76**, 1256–1265 (2019).

21.	Lee, J. J. *et al.* Gene discovery and polygenic prediction from a genome-wide association study of educational attainment in 1.1 million individuals. *Nature Genetics* **50**, 1112–1121 (2018).

22.	Yengo, L. *et al.* Meta-analysis of genome-wide association studies for height and body mass index in ∼700000 individuals of European ancestry. *Hum Mol Genet* **27**, 3641–3649 (2018).

23.	Lu, Q. *et al.* A Powerful Approach to Estimating Annotation-Stratified Genetic Covariance via GWAS Summary Statistics. *Am J Hum Genet* **101**, 939–964 (2017).

24.	Davey Smith, G. & Hemani, G. Mendelian randomization: genetic anchors for causal inference in epidemiological studies. *Hum. Mol. Genet.* **23**, R89-98 (2014).

25.	Davies, N. M., Holmes, M. V. & Davey Smith, G. Reading Mendelian randomisation studies: a guide, glossary, and checklist for clinicians. *BMJ* **362**, k601 (2018).

26.	Morrison, J., Knoblauch, N., Marcus, J. H., Stephens, M. & He, X. Mendelian randomization accounting for correlated and uncorrelated pleiotropic effects using genome-wide summary statistics. *Nat. Genet.* **52**, 740–747 (2020).

27.	Hemani, G. *et al.* The MR-Base platform supports systematic causal inference across the human phenome. *eLife* **7**, e34408 (2018).

28.	Ruth Mitchell, E. MRC IEU UK Biobank GWAS pipeline version 2. *data.bris* https://data.bris.ac.uk/data/dataset/pnoat8cxo0u52p6ynfaekeigi (2019) doi:10.5523/bris.pnoat8cxo0u52p6ynfaekeigi.

29.	Bowden, J., Davey Smith, G., Haycock, P. C. & Burgess, S. Consistent Estimation in Mendelian Randomization with Some Invalid Instruments Using a Weighted Median Estimator. *Genet. Epidemiol.* **40**, 304–314 (2016).

30.	Bowden, J., Davey Smith, G. & Burgess, S. Mendelian randomization with invalid instruments: effect estimation and bias detection through Egger regression. *Int J Epidemiol* **44**, 512–525 (2015).

31.	Hartwig, F. P., Davey Smith, G. & Bowden, J. Robust inference in summary data Mendelian randomization via the zero modal pleiotropy assumption. *Int J Epidemiol* **46**, 1985–1998 (2017).

32.	Verbanck, M., Chen, C.-Y., Neale, B. & Do, R. Detection of widespread horizontal pleiotropy in causal relationships inferred from Mendelian randomization between complex traits and diseases. *Nat. Genet.* **50**, 693–698 (2018).

33.	Bowden, J. *et al.* Improving the visualization, interpretation and analysis of two-sample summary data Mendelian randomization via the Radial plot and Radial regression. *Int J Epidemiol* **47**, 1264–1278 (2018).
