## Supplementary Figures for "Unraveling the relationship of loneliness and isolation in schizophrenia: Polygenic dissection and causal inference"

**Table of Contents**

**Supplementary Figure 1. PGS_LNL-ISO_ predictions after 10,000 permutations of the SCZ - HC status …………………………............................................................................................................. 2**

**Supplementary Figure 2. PGS predictions after 10,000 permutations of the SCZ - HC status for LNL-ISO related traits from UKBB …………………………………………………………..…….... 3**

**Supplementary Figure 3. Beta sign concordance between SCZ and LNL-ISO across different SCZ p-thresholds …………………………………………………………………………………………... 4**

**Supplementary Figure 4. Results from the partitioned heritability analysis with LDSR for tissue and cell-type enrichment ……………..………………………………………………………….. 5**

**Supplementary Figure 5. Results from Mendelian Randomization with IVW and WM methods and sensitivity test of Loneliness and isolation traits and Schizophrenia liability ……………..……………………………………………………...…………………………………………… 6**

**Supplementary Figure 6. Single-SNP effect and leave-one-out sensitivity tests of Mendelian Randomization analyses of LNL-ISO and Loneliness UKBB on schizophrenia liability………………………...……………………………………………...………………………………. 7**

**Supplementary figure 1.** **PGS_LNL-ISO_ predictions after 10,000 permutations of the SCZ - HC status**. Predicted pseudo-R^2^ was compared to the Naggelkerke’s pseudo-R^2^ distribution after 10,000 permutations.

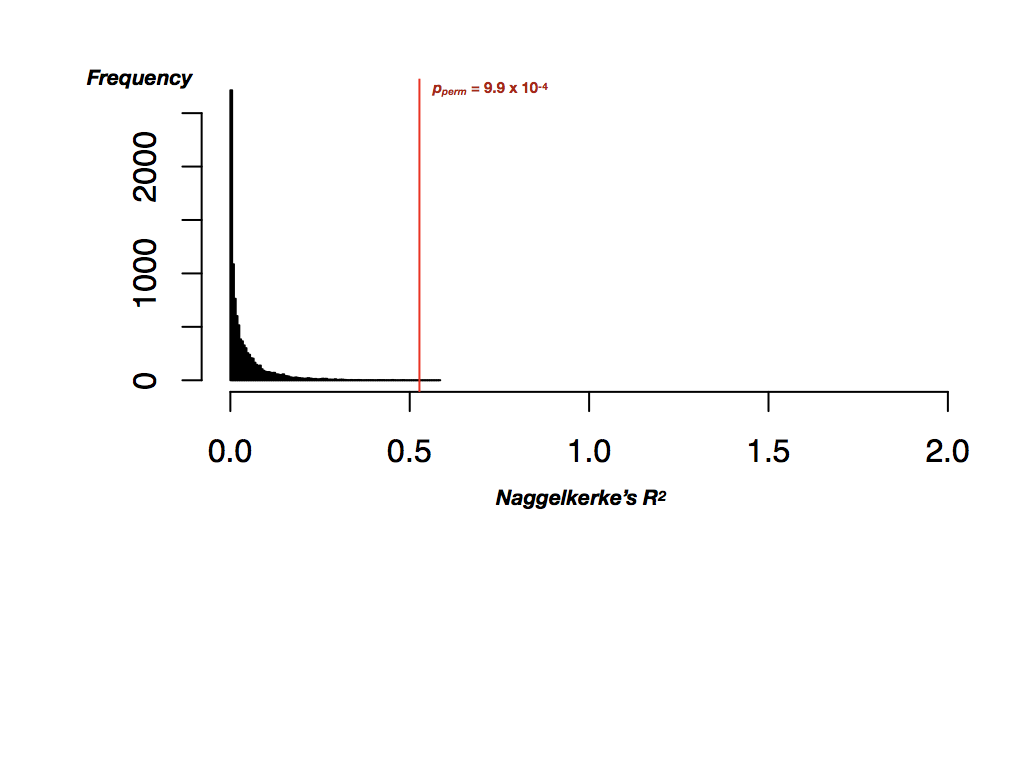

**Supplementary figure 2.** **PGS predictions after 10,000 permutations of the SCZ - HC status for LNL-ISO constituent traits from UKBB**. Predicted pseudo-R^2^ was compared to the Naggelkerke’s pseudo-R^2^ distribution after 10000 permutations for A) loneliness UKBB (<https://nealelab.github.io/UKBB_ldsc/h2_summary_2020.html>), B) number of family/friends visits (<https://nealelab.github.io/UKBB_ldsc/h2_summary_1031.html>), C) number of people in the household (<https://nealelab.github.io/UKBB_ldsc/h2_summary_709.html>) and D) ability to confide in others ([https://nealelab.github.io/UKBB_ldsc/h2_summary_2110.html](https://nealelab.github.io/UKBB_ldsc/h2_summary_2020.html)).

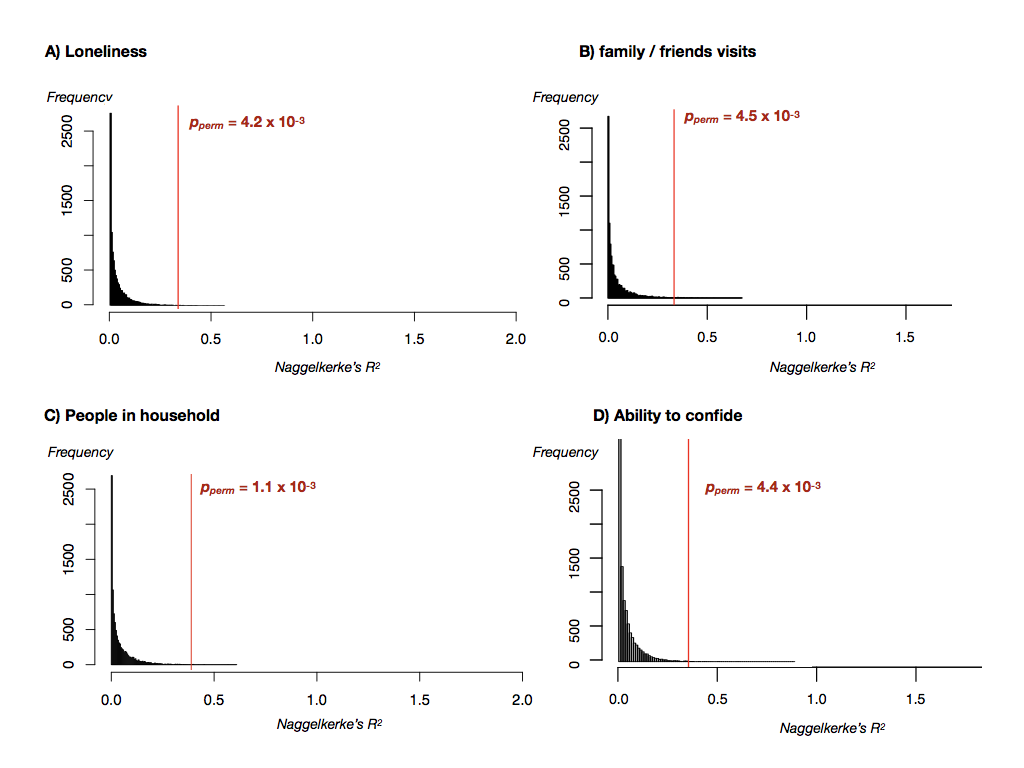

**Supplementary Figure 3. Results from the partitioned heritability analysis with LDSR for tissue and cell-type enrichment.** Green, red and blue bars represent heritability enrichment for 10 tissues (Bullik-Sullivan et al., 2015; http://data.broadinstitute.org/alkesgr oup/LDSCORE/), 13 brain tissues from GTEx (Bullik-Sullivan et al., 2015; <http://data.broadinstitute.org/alkesgroup/LDSCORE/>) and 3 brain cell-types from Cahoy et al. (2008) (Finucane et al., 2018; http://data.broadinstitute.org/alkesgroup/LDSCORE/) across i) SCZ_noLNL (i.e., SNPs from SCZ GWAS not associated with LNL-ISO (P > 0.05)) and ii) SCZ_LNL_CONC (i.e., SNPs from SCZ GWAS associated with LNL-ISO (P < 0.05)) genome annotations. In brain tissue and cell type graphs, target tissue is compared to ‘anti-target tissue’ enrichment (see Supplementary Methods). ***** significant results in the comparison between target and anti-target tissue enrichment after Benjamini-Hochberg FDR correction. + significant results at the non-corrected level in the target tissue vs anti-target tissue enrichment comparison. Dashed lines and shading around them represent the enrichment and its standard deviation for the control tissue in each case. ACC: Anterior cingulate cortex; CNS: Central nervous system

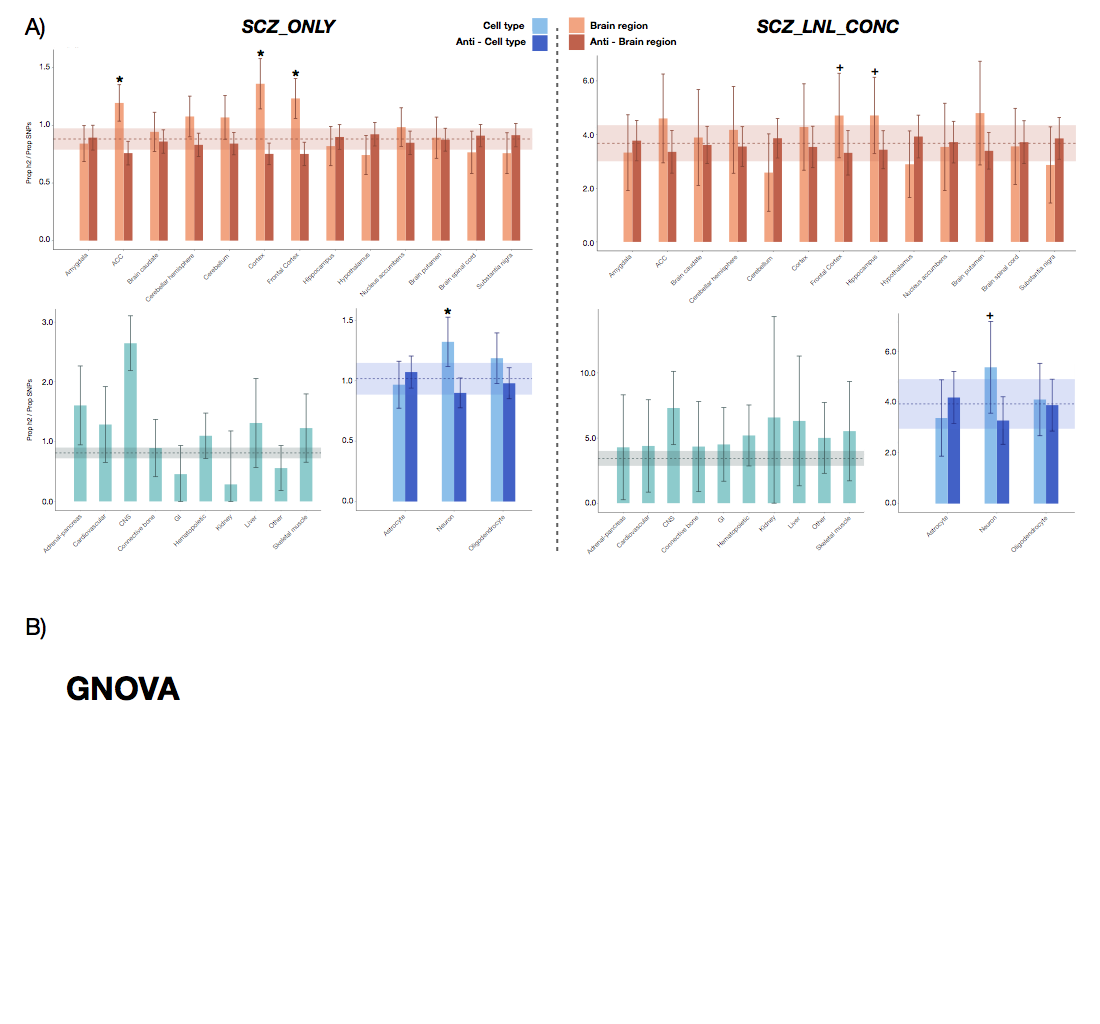

**Supplementary Figure 4. Single-SNP effect and Leave-one-out Sensitivity test of Mendelian Randomization analyses of LNL-ISO and Loneliness UKBB on Schizophrenia liability. A** Forest Plots of Single SNP effects(left) and Leave-one-out analysis (right) of LNL-ISO against Schizophrenia liability. **B** Forest Plots of Single SNP effects(left) and Leave-one-out analysis (right) of Loneliness UKBB against Schizophrenia liability. Note that a single variable (rs159960) IN Loneliness UKBB causes a greater effect than the rest of the instruments in the leave-one-out test and has a discordant effect relative to the rest of the traits.

**A**

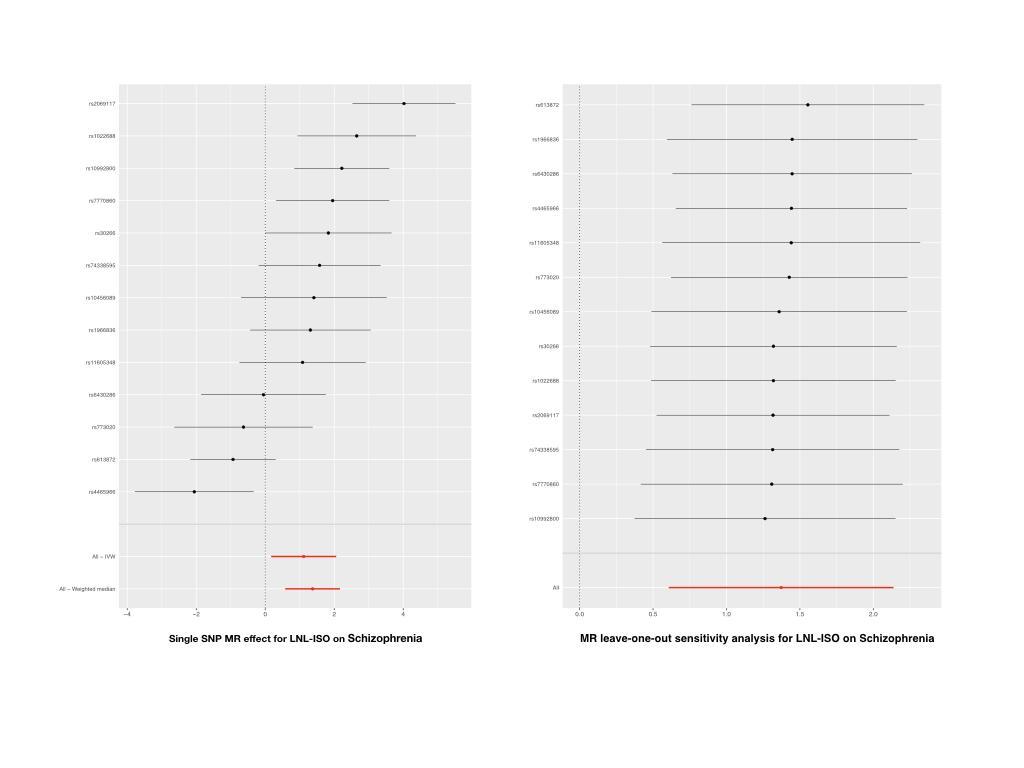

**B**

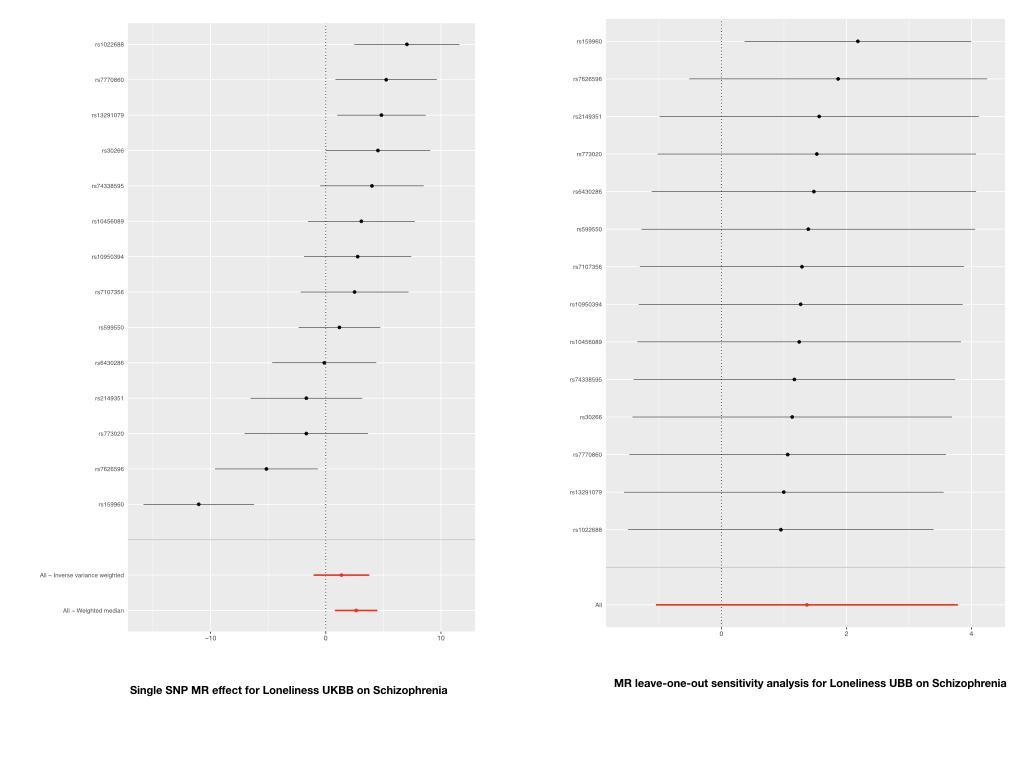

**Supplementary Figure 5. Results from Bidirectional Mendelian Randomization with IVW and WM methods of loneliness and isolation traits and schizophrenia liability.** One-to-many forest plots of the results of bidirectional causality analysis with two MR methods. Red dots represent a significant positive causal association. Blue dots represent a negative or protective causal association

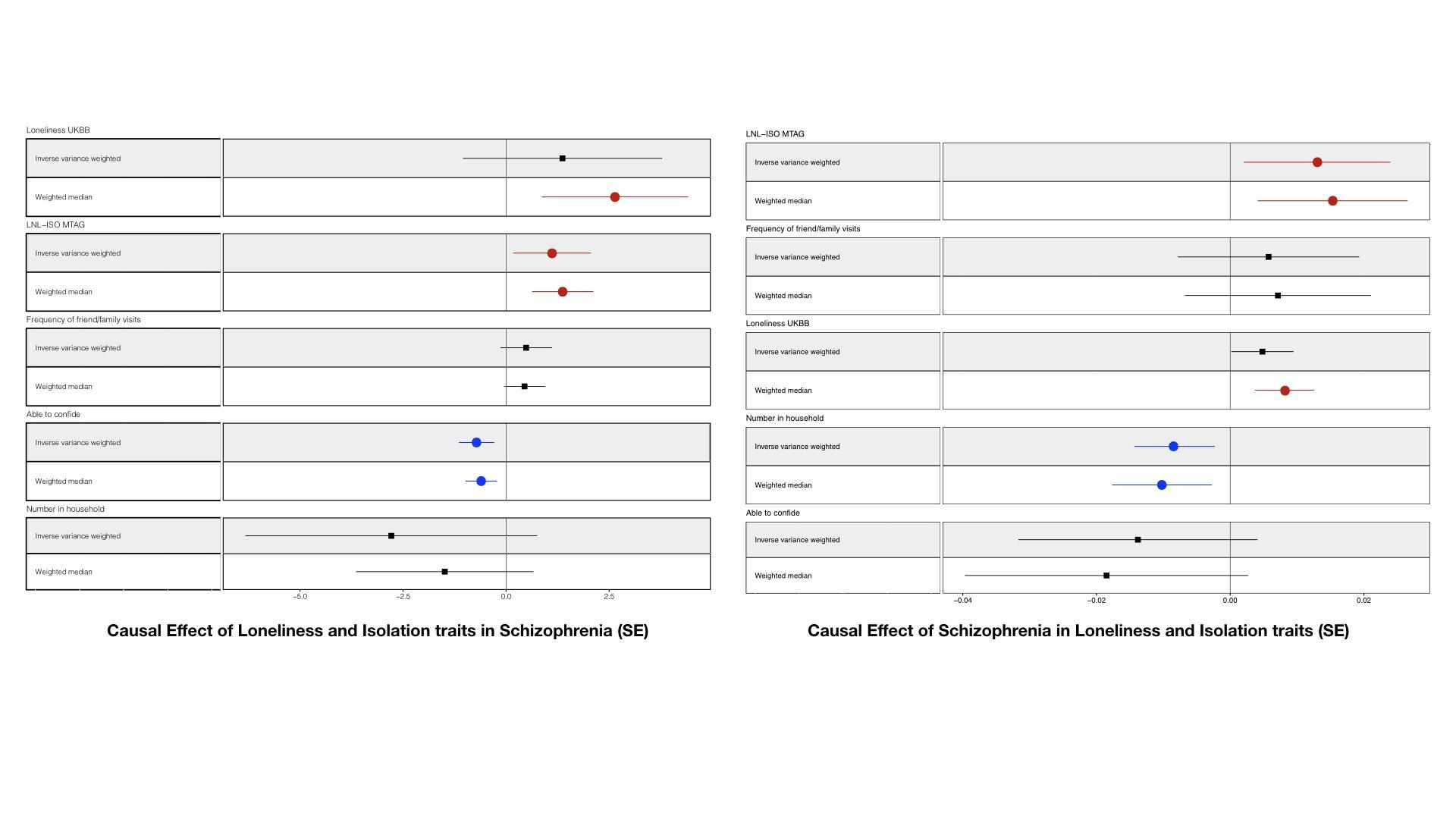
